# First in Human study of a microRNA29a mimic (TenoMiR) in patients with lateral elbow tendinopathy - a randomised, Placebo Controlled Phase 1 trial

**DOI:** 10.1101/2025.08.23.25332854

**Authors:** Neal L Millar, Iona Donnelly, Iain B McInnes, David Ball, Pui Man Leung, Annad Kirwadi, D. S. Gilchrist

## Abstract

Tendinopathy, encompassing multifactorial tendon disorders characterised by pain and functional limitation, remains a significant burden in musculoskeletal medicine. Here we report a prospective, randomised, double-blind, placebo-controlled Phase 1b first in patient trial of three single ascending doses of intratendinous injection of TenoMiR, a chemically synthesised mimic of microRNA-29a versus placebo conducted in 24 subjects with active lateral elbow tendinopathy (NCT04670289). The primary objective of the study was to determine the safety and tolerability of single ascending doses of TenoMiR in subjects with lateral epicondylitis, with patient reported measures and imaging parameters as secondary outcomes. TenoMiR treatment at 200, 500 or 1500 µg/mL was associated with acceptable safety and tolerability with the geometric mean maximum observed value (Cmax,relative expression compared to control) of miR29a was 3.16, 6.63 and 2.37 respectively. There were no significant treatment- or dose-related trends in clinical laboratory evaluations, vital signs or ECG parameters. Treatment with TenoMiR led to a substantial improvement in tendon structure at all doses over a 90 day period and there was improvement of symptoms and function in both groups. This data supports the mechanism of miRNA-mediated modulation of the early pathophysiologic events that facilitate tissue remodelling in human tendon disease, and provides strong proof of principle that a locally delivered miR29a therapy improves early tendon healing.

## Introduction

Overuse tendon injuries, namely tendinopathies, pose a significant, highly prevalent problem in musculoskeletal medicine. The diagnosis and management of shoulder tendon injuries alone amounts to an annual cost of $3 billion to the US healthcare system highlighting the huge burden of disease.^1^ The paucity of effective treatments in tendon disorders partially reflects lack of understanding of its pathogenesis.^2^ The immune system plays a crucial role in the regulation of tissue remodelling by coordinating complex signalling networks that facilitate transcriptional regulation of extracellular matrix (ECM) components as an adaptive response to environmental cues.^3^ Inflammatory mediators are considered crucial to the onset and perpetuation of tendinopathy.^2,4,5^ Expression of various cytokines has been demonstrated in inflammatory cell lineages and tenocytes, suggesting that both infiltrating and resident populations participate in pathology.^6–8^ Additionally, evidence suggests a balance exists between pro-inflammatory and pro-resolving mediators that may ultimately define the extent to which tendinopathy develops and subsequently repairs.^9,10^ Recent work has identified the importance of tissue microenvironments and the interaction immune mediators in inflammatory/stromal cell crosstalk.^11^

MicroRNAs (miRNAs) are small, non-coding RNAs that suppress gene expression at the post-transcriptional level by inhibiting translation and/or inducing mRNA degradation.^12^ A single miRNA can regulate the expression of multiple genes through binding of complementary sequences in the 3’ untranslated region of target mRNAs. This property renders miRNAs attractive as potential therapeutic tools to restore cell functions that are altered as part of a disease phenotype. MiRNAs have emerged that particularly regulate cytokine networks while orchestrating proliferation and differentiation of stromal lineages that determine ECM composition.^13^ miRNAs have provoked extensive interest as regulators of musculoskeletal diseases, although their precise contributions to complex disease pathways remains uncertain.^14^

We have previously demonstrated a functional role for miR-29a as a post-transcriptional regulator of collagen in murine, equine and human tendon injury^15^. In healthy tenocytes, miR-29a directly suppresses collagen 3 over-production, neo-vascularisation, hyper-proliferation of tenocytes and by targeting Col3a1, vascular endothelial growth factor A (VEGFA), AKT3 and TGF-β2, respectively. Significantly, the sequence of miR-29a and its binding sites in these target genes are conserved in mammalian species. Chemical modifications offer a possibility to enhance the cellular uptake of oligonucleotides. Modifications of the phosphate backbone, the nucleobases, or the ribose sugar can mask the charge of the miRNAs and further their adhesion to the cell surface, thereby facilitating the cellular uptake. TenoMiR is a chemically synthesised mimic of miR29a which has improved stability, activity and cellular uptake, while being non-immunogenic and has been created to restore miR29a functional expression back to homeostatic levels in tendon. The sequence of the antisense strand of TenoMiR is identical to natural miR29a and therefore inhibits expression of the same target genes. To increase stability and activity, TenoMiR’s backbone has been chemically modified via the introduction of 2’Fluro and 2’O-methyl groups, and the sense strand contains a 3’ cholesterol group to increase cellular uptake. Given the translational potential we designed a Phase I First in Human study to evaluate the potential for miR-29a replacement therapy as a therapeutic option to treat tendon disease in patients with lateral elbow tendinopathy.

## Methods

### Study design and participants

This was a Phase 1, single site, randomised, double-blind, placebo-controlled study evaluating the safety, tolerability and PK of single ascending doses of TenoMiR injections in subjects with lateral epicondylitis (see Figure 1 for study design). Subjects were divided into cohorts (8 subjects per cohort) and were randomised (3:1) to receive a single 1 mL dose of TenoMiR or saline placebo (0.9%) administered via injection into the tendon lesion. The starting dose of TenoMiR was 200 µg/mL escalating to 500 µg/mL and then 1500 µg/mL. All doses were given as a 1 mL injection under ultrasound guidance into the affected area. In each cohort, no more than 2 subjects on the first dosing day (1 active; 1 placebo) were dosed, such that no more than 1 subject received an active TenoMiR dose for the first time at each dose level. After 72 hours, depending on the safety and tolerability of the previously dosed sentinel subjects, dosing continued in the remaining subjects (6 subjects; 5 active, 1 placebo) at the same dose level in each cohort. Doses were administered in an escalating manner, following satisfactory review of all safety and tolerability data from lower doses. Except for the starting dose, the dose levels studied were preliminary, with actual subsequent doses determined based on ongoing evaluations of the safety, tolerability, plasma PK and efficacy data (where available) by the safety review committee (SRC). A copy of the full trial protocol is available at https://clinicaltrials.gov/study/NCT04670289.

**Figure 1.**
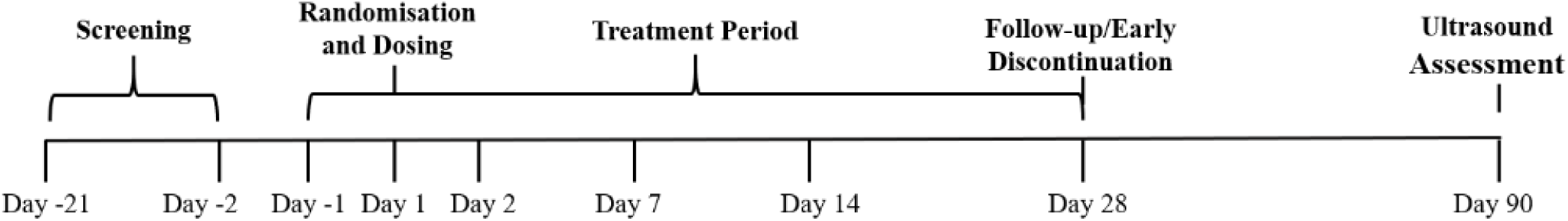
Study design.

The primary endpoint of the study was the comparison of safety data between TenoMiR versus placebo as measured by incidence of adverse events (AEs), clinical laboratory abnormalities, changes in vital signs (blood pressure, temperature, respiratory rate and pulse rate), 12-lead electrocardiogram (ECG) parameters, physical examinations and skin score assessment at 14 days post-injection. The secondary endpoints of the study were the plasma pharmacokinetics of TenoMiR (C_max_, C_max_ (t_max)_ and AUC), the efficacy of a single dose of TenoMiR on elbow pain as measured using VAS, on disability and symptoms measured using the Disabilities of the Arm, Shoulder and Hand (Quick-DASH), on pain and disability as measured by the American Shoulder and Elbow Surgeons Elbow (ASES-E) Score, on pain and disability as measured by the Patient Rated Tennis Elbow Evaluation (PRTEE) and Ultrasound Tissue Characterisation imaging from predose to Day 14, Day 28 and Day 90.

### Blinding

This study was double-blinded (Investigator and subject-blinded). After informed consent was obtained, subjects were allocated a unique Screening number. Only subjects who complied with all the inclusion criteria and none of the exclusion criteria, were randomised onto the study. The subjects were assigned to a randomisation number in the order of recruitment. All screened subjects were identifiable throughout the study. A randomisation list, detailing which cohort the subject was allocated to, was generated using SAS PROC PLAN. The randomisation list was kept in a secure location until the end of the study. Only the Pharmacy staff involved in handling the study drug and the laboratory staff responsible for analysing the PK blood samples were unblinded during the study and had access to the randomisation list. The planned volume of either TenoMiR or placebo were given as identical syringes to the dosing staff in the clinical research site.

### Ultrasound Assessment

Recently, a novel imaging modality called Ultrasound Tissue Characterisation (UTC) ^16^ has been developed to visualise tendon structure and to quantify tendon matrix integrity. Unlike two-dimensional ultrasound and colour Doppler, UTC objectively quantified grey-scale tendon matrix changes into 4 different echotypes related to tendon integrity. Types I (green) and II (blue) represent an organised matrix; Types III (red) and IV (black) represent a disorganised matrix. The advantage of this tool, compared to conventional ultrasound, is that it captures a three-dimensional ultrasound image of the tendon and uses standardised parameters (transducer tilt angle, depth and gain settings). A scan of the affected elbow was performed by two examiners with experience performing UTC scans. Images were acquired using a 7-10 MHz linear ultrasound transducer (SmartProbe 12L5-V, Terason 2000+; Teratech; Burlington, MD, USA) positioned in a tracking device (UTC Tracker, UTC Imaging, Stein, The Netherlands) that scanned the tendon’s long axis over a distance of 5 cm recording regular images at intervals of 0.2 mm. UTC takes 647 images along the forearm that includes the key section of the common extensor origin tendon. Transducer tilt, angle, gain, focus and depth are standardised by the tracking device. Images from the sagittal, coronal and transverse planes are compiled to create a three-dimensional view of the tendon. UTC assessment were conducted pre-treatment (within 4 weeks) and at Days 28 and 90 post-treatment. The images over the common extensor tendon were then reviewed in both the sagittal and coronal plane, and three points along the length of the core extensor tendon which are 0.5 cm starting from the lateral epicondyle. A ellipse area of a 40 × 40 mm (the maximal tendon thickness is estimated in humans to be 45 mm) was applied at 0.5,1 and 1.5 cm from the lateral epicondyle and at each point provided percentage scores of echo types I-IV thus giving standardised scoring across the whole tendon for the patient population. Images were reviewed blind by a Consultant Orthopaedic Surgeon and Consultant Radiologist trained in UTC, and the mean area score from each scorer gave the final value displayed in the following figures. Six blinded subject images (total of 18 separate UTC images) were used to determine the inter-rater reliability of r = 0.87.

### Statistical and Analytical Plans

The planned statistical methodology for the analysis of the safety, PK and efficacy data is detailed in the Statistical Analysis Plan (SAP) and PK SAP (Available on request). 24 subjects with lateral epicondylitis were enrolled in this study. Given the exploratory nature of this study, the sample size was not based on power calculations. The number of subjects included is typical for this kind of study and the results provide sufficient safety and tolerability data, in addition to adequate description of the PK and efficacy parameters, without exposing too many subjects. The inclusion of placebo subjects permitted informal comparisons between the TenoMiR treated subjects and of each dose cohort versus the complete placebo cohort. The effects of TenoMiR on efficacy parameters was analysed with a Generalised Linear Mixed Model (GLMM) using an efficacy parameter as responsible variable, treatment (active and placebo), timepoint, interaction between treatment and timepoint as mixed effects, baseline measurements as a covariate, subject and elbow (left and right) as random effects. This model was applied to each efficacy parameter. Treatment differences between active and placebo at different timepoints together with 95% CIs were derived from the generalised linear mixed model. For a continuous outcome, treatment difference was measured as the difference in least square means; for a binary outcome, treatment difference was measured as the odds ratio.

### Patients

Male and female subjects aged 18–70 years fulfilling the following criteria were eligible for enrolment: unilateral lateral epicondylitis (LE) tendinopathy with symptoms present for at least 6 weeks but not more than 6 months. The tendinopathy must have been refractory to standard treatment (NSAIDs/paracetamol, physiotherapy, and/or steroid injection). In the run-in period, the patient stopped NSAID intake for 1 week prior to injection. Key inclusion criteria included LE tendinopathy with symptoms that could be reproduced with resisted supination or wrist dorsiflexion (as confirmed by tenderness at lateral epicondyle and positive pick up back of chair sign). If the subject had previous steroid treatment, it must have been > 6 months weeks prior to randomisation. Key exclusion criteria and considerations regarding concomitant medications are summarised in the supplemental material.

### Patient and public involvement

Patients or the public were not involved in the design and conduct of the trial. The trial was conducted in accordance with the Declaration of Helsinki (General Assembly of the World Medical Association 2014) and was approved by the Independent Ethics Committee or Institutional Review Board for the centre. Written informed consent was obtained from all enrolled patients. Data were collected in accordance with Good Clinical Practice guidelines by the trial investigators and analysed by MAC Clinical Research.

## Results

### Demographic Data

Twenty-four subjects were enrolled, randomised evenly across three dose cohorts and dosed during the study. All 24 subjects completed the study. There were 16 (66.7%) male subjects and 8 (33.3%) female subjects enrolled onto this study. All placebo subjects were male. The majority of subjects (91.7%) were white. Mean age was generally similar across the active treatment groups but the mean age in all three placebo groups was higher than the overall mean. The mean weight in the 500 µg/mL TenoMiR treatment group was lower than the overall mean and the mean weights in the three placebo treatment groups were higher than those of subjects in the corresponding active group. The mean height and BMI were generally similar across treatment groups ^17^. Demographic data is summarised in Table 1 with identifying information removed.

**Table 1.**
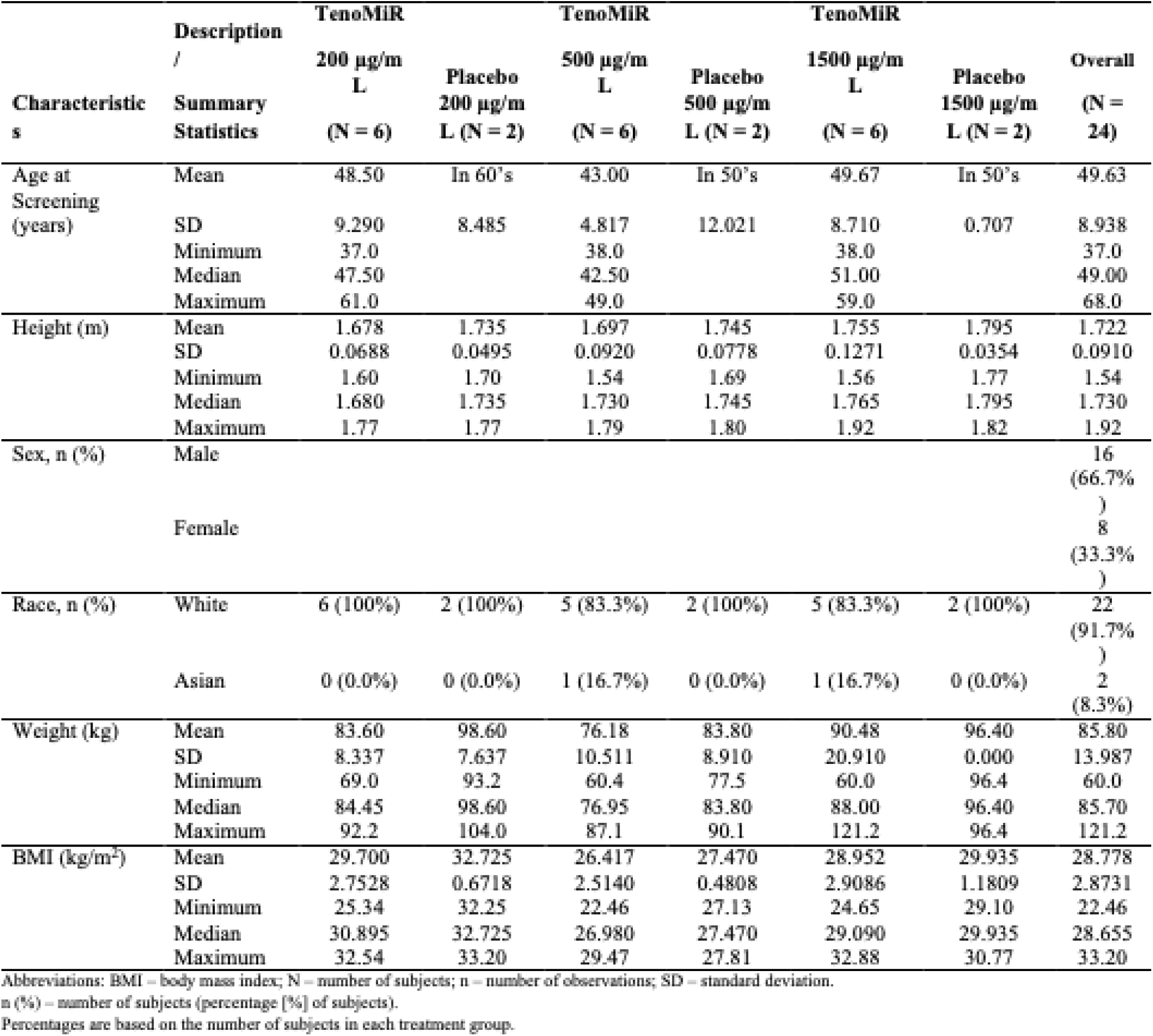
Patient demographics.

**Table 2.** Change from baseline in Visual Analogue Pain scale.

**Change from Baseline in Elbow Pain/Tendon Pain Questionnaire (VAS) Scores by GLMM Analysis, Multi-Item Symptom Scales**
| Visit | Statistic | TenoMiR<br>200 µg/mL<br>(N=6) | Placebo<br>200 µg/mL<br>(N=2) | TenoMiR<br>500 µg/mL<br>(N=6) | Placebo<br>500 µg/mL<br>(N=2) | TenoMiR<br>1500 µg/mL<br>(N=6) | Placebo<br>1500 µg/mL<br>(N=2) |
| --- | --- | --- | --- | --- | --- | --- | --- |
| Day 14 | n | 6 | 2 | 6 | 2 | 6 | 2 |
|  | Adjusted mean [a] | -19.0 | -42.0 | -13.9 | -4.2 | -20.3 | -24.7 |
|  | Standard Error | 9.40 | 16.48 | 5.89 | 10.47 | 9.18 | 16.14 |
|  | 95% CI | -38.10, 0.18 | -75.53, -8.41 | -25.90, -1.94 | -25.51, 17.03 | -38.90, -1.60 | -57.56, 8.06 |
|  | Estimated difference [b] | 23.0 |  | -9.7 |  | 4.5 |  |
|  | 95% CI for difference | -15.84, 61.85 |  | -34.54, 15.18 |  | -33.68, 42.67 |  |
|  | p-value | 0.2365 |  | 0.4342 |  | 0.8122 |  |
| Day 28<br>(FU) | n | 5 | 2 | 6 | 2 | 6 | 2 |
|  | Adjusted mean [a] | -21.2 | -44.0 | -21.9 | -27.2 | -11.8 | -30.7 |
|  | Standard Error | 9.89 | 16.48 | 5.89 | 10.47 | 9.18 | 16.14 |
|  | 95% CI | -41.32, -1.03 | -77.53, -10.41 | -33.90, -9.94 | -48.51, -5.97 | -30.48, 6.81 | -63.56, 2.06 |
|  | Estimated difference [b] | 22.8 |  | 5.3 |  | 18.9 |  |
|  | 95% CI for difference | -16.60, 62.19 |  | -19.54, 30.18 |  | -19.26, 57.09 |  |
|  | p-value | 0.2473 |  | 0.6665 |  | 0.3211 |  |
| Day 90<br>(EOS) | n | 6 | 2 | 6 | 2 | 6 | 2 |
|  | Adjusted mean [a] | -36.8 | -47.5 | -36.5 | -31.2 | -26.1 | -31.7 |
|  | Standard Error | 9.40 | 16.48 | 5.89 | 10.47 | 9.18 | 16.14 |
|  | 95% CI | -55.94, -17.65 | -81.03, -13.91 | -48.48, -24.53 | -52.51, -9.97 | -44.73, -7.44 | -64.56, 1.06 |
|  | Estimated difference [b] | 10.7 |  | -5.3 |  | 5.7 |  |
|  | 95% CI for difference | -28.17, 49.51 |  | -30.13, 19.60 |  | -32.51, 43.84 |  |
|  | p-value | 0.5796 |  | 0.6696 |  | 0.7648 |  |
| Average | n | 6 | 2 | 6 | 2 | 6 | 2 |
|  | Adjusted mean [a] | -25.9 | -44.5 | -24.1 | -20.9 | -19.4 | -29.1 |
|  | Standard Error | 7.98 | 13.96 | 4.48 | 8.10 | 6.39 | 11.43 |
|  | 95% CI | -42.17, -9.64 | -72.91, -16.03 | -33.23, -15.00 | -37.37, -4.44 | -32.38, -6.40 | -52.31, -5.86 |

**Table 3.** Overall summary of treatment emergent adverse events.

Overall Summary of Treatment-Emergent Adverse Events
|  | **TenoMiR**<br>**200 µg/mL**<br>**(N = 6)** | **Placebo**<br>**200 µg/mL**<br>**(N = 2)** | **TenoMiR**<br>**500 µg/mL**<br>**(N = 6)** | **Placebo**<br>**500 µg/mL**<br>**(N = 2)** | **Ten**<br>**1500**<br>**(N** |
| --- | --- | --- | --- | --- | --- |
| Any TEAE | 4 (66.7%), 16 | 2 (100%), 8 | 6 (100%), 26 | 2 (100%), 2 | 3 (50. |
| Any Serious TEAE | 1 (16.7%), 1 | 0 (0.0%), 0 | 0 (0.0%), 0 | 0 (0.0%), 0 | 0 (0.) |
| Any TEAE Leading to<br>Discontinuation of<br>Treatment | 0 (0.0%), 0 | 0 (0.0%), 0 | 0 (0.0%), 0 | 0 (0.0%), 0 | 0 (0.) |
| Due to Study<br>Medication-Related<br>TEAE | 0 (0.0%), 0 | 0 (0.0%), 0 | 0 (0.0%), 0 | 0 (0.0%), 0 | 0 (0.) |
| Due to Serious<br>TEAE | 0 (0.0%), 0 | 0 (0.0%), 0 | 0 (0.0%), 0 | 0 (0.0%), 0 | 0 (0.) |
| Other Reason | 0 (0.0%), 0 | 0 (0.0%), 0 | 0 (0.0%), 0 | 0 (0.0%), 0 | 0 (0.) |
| Any Life-Threatening<br>SAEs | 1 (16.7%), 1 | 0 (0.0%), 0 | 0 (0.0%), 0 | 0 (0.0%), 0 | 0 (0.) |
| TEAE Leading to<br>Death | 0 (0.0%), 0 | 0 (0.0%), 0 | 0 (0.0%), 0 | 0 (0.0%), 0 | 0 (0.) |
| Severity |  |  |  |  |  |
| Mild | 4 (66.7%), 11 | 2 (100%), 6 | 6 (100%), 22 | 2 (100%), 2 | 3 (50. |
| Moderate | 1 (16.7%), 1 | 1 (50.0%), 2 | 3 (50.0%), 4 | 0 (0.0%), 0 | 0 (0.) |
| Severe | 1 (16.7%), 4 | 0 (0.0%), 0 | 0 (0.0%), 0 | 0 (0.0%), 0 | 0 (0.) |
| Causality (All TEAEs) |  |  |  |  |  |
| Not Related | 4 (66.7%), 8 | 2 (100%), 4 | 6 (100%), 15 | 1 (50.0%), 1 | 3 (50. |
| Unlikely Related | 2 (33.3%), 7 | 2 (100%), 2 | 4 (66.7%), 7 | 0 (0.0%), 0 | 0 (0.) |
| Possibly Related | 1 (16.7%), 1 | 1 (50.0%), 2 | 2 (33.3%), 2 | 1 (50.0%), 1 | 0 (0.) |
| Probably Related | 0 (0.0%), 0 | 0 (0.0%), 0 | 2 (33.3%), 2 | 0 (0.0%), 0 | 0 (0.) |
| Related [a] | 1 (16.7%), 1 | 1 (50.0%), 2 | 3 (50.0%), 4 | 1 (50.0%), 1 | 0 (0.) |

Abbreviations: N – number of subjects; SAE – serious adverse event; TEAE – treatment-emergent adverse event.
xx (xx.x), xx: number of subjects (percentage of subjects), number of events.
Percentages (xx.x) are based on the total number of subjects in each treatment group.
A TEAE is defined as an adverse event with a start date on or after dosing with TenoMiR/placebo.
[a] Treatment-related = relationship to study drug is answered 'Probably Related' or 'Possibly Related'

### Pharmacokinetics

Following a single injection of TenoMiR at 200, 500 or 1500 µg/mL, the geometric mean maximum observed value (Cmax) (PK was expressed as fold change over baseline) of miR29a was 3.16, 6.63 and 2.37 respectively. Following a single injection of placebo, the geometric mean Cmax value of miR29a was 1.90 and was attained on average (median tmax) at 7 hours postdose, with individual tmax estimates observed from baseline to 24 hours postdose. Systemic exposure to miR29a (Cmax and AUC0-t) tended to decrease with dose; however, there was no apparent dose proportional relationship. The Cmin values decreased with dose and the change was less than dose proportional. For a doubling in dose, Cmin was predicted to decrease 1.6 fold and the 90% CIs (1.22, 2.07) were outside the prescribed limits of 1.6 to 2.5; therefore, dose proportionality could not be confirmed. A full PK report is available upon request.

### Safety

Single injections of TenoMiR appeared to be safe and well tolerated when administered over a dose range of 200 to 1500 µg/mL, with no dose-related trend observed in the frequency of TEAEs. No deaths or TEAEs leading to withdrawal occurred in the study. One SAE of a suspected COVID-19 infection that required hospitalisation was reported by 1 subject in the 200 µg/mL TenoMiR treatment group. Overall, 18 (75.0%) subjects experienced 59 TEAEs across all treatment groups. All 6 subjects in the 500 µg/mL TenoMiR treatment group, and both subjects in each of the 200 µg/mL placebo and 500 µg/mL placebo groups reported at least one TEAE. Of the 59 TEAEs reported, 48 events reported by 18 (75.0%) subjects were mild in severity, 7 events reported by 5 (20.8%) subjects were moderate and 4 events reported by 1 (4.2%) subject were severe. All 4 severe events were reported by 1 subject in the 200 µg/mL TenoMiR treatment group and were not considered to be related to treatment. Eight of the 59 reported TEAEs were considered to be possibly or probably related to treatment. Of these 8 events, 5 were reported by subjects administered TenoMiR and 3 by subjects administered placebo. There were no treatment related TEAEs reported at the highest dose level (1500 µg/mL TenoMiR) or in the 1500 µg/mL placebo group. The most commonly reported TEAEs were within the musculoskeletal and connective tissue disorders and nervous system disorders SOCs. Within these SOCs, the most common TEAEs (by PT) were headache, arthralgia, joint stiffness and pain in extremity. With the exception of contusion, all other TEAEs were reported by a single subject. There were no significant treatment- or dose-related trends in the mean or individual subject haematology, serum biochemistry or urinalysis data during the study. There were no significant treatment- or dose-related trends in the mean or individual subject vital sign values over the 200 to 1500 µg/mL TenoMiR dose range or placebo for systolic blood pressure, diastolic blood pressure, pulse rate, respiratory rate or oral temperature. Although some vital sign parameters were outside appropriate reference ranges, these findings were observed at an isolated timepoint, were transient and held no clinical significance. There were no significant treatment- or dose-related trends in the mean ECG parameters over the 200 to 1500 µg/mL TenoMiR dose range or placebo. Although some ECG parameters were outside appropriate reference ranges at some timepoints, these findings were determined to be transient and of no clinical significance.

### Clinical Efficacy

#### Key pain and function outcomes (secondary endpoints) Visual analogue pain score

Decreases from baseline in mean pain/tendon pain questionnaire VAS scores were observed for all active treatment groups and all placebo groups at all the assessed post-injection timepoints. There was no apparent dose-related trend in the decreases from baseline for the active groups. There were no statistically significant differences in mean change from baseline in elbow pain/tendon pain questionnaire (VAS) scores for all TenoMiR dose groups compared to placebo (Figure 2A-C). In the total study population, treatment with TenoMiR resulted in clinically relevant improvements in VAS score at Days 14,28 and 90 compared to placebo which had clinically relevant improvements at Days 14 and 90 (Figure 2D).

**Figure 2.**
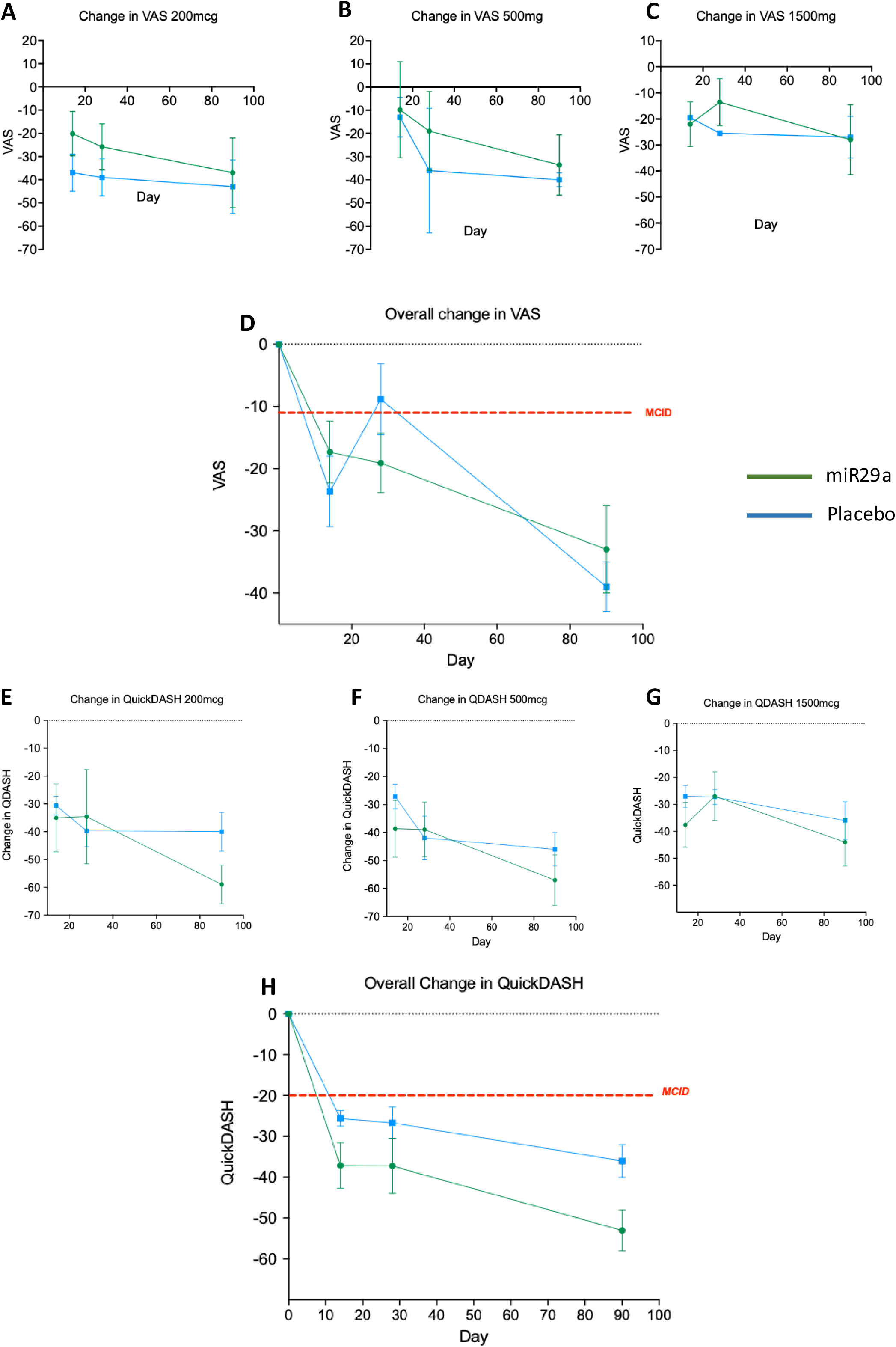
Visual analogue pain score and QuickDASH patient reported outcomes. (A-D) change in pain (VAS) score in 200mcg,500mcg, 1500mcg (n=6 miR29a, n=2 placebo) and overall patient population (n=18 miR29a, n=6 placebo) and (E-H) change in QuickDASH total score in 200mcg,500mcg, 1500mcg (n=6 miR29a, n=2 placebo) and overall patient population (n=18 miR29a, n=6 placebo) in the microRNA29a mimic and placebo groups through Day 90, adjusted means (SE); VAS, visual analogue scale; miR29a, microRNA 29a, QDASH, Quick Disabilities of Arm,Shoulder and Hand; MCID, Minimum Clinically Important Difference

#### Patient-reported outcomes: QuickDASH, ASES-E, PRTEE

Decreases from baseline in mean Quick-DASH Disability/Symptom Scores were observed for all active treatment groups and all placebo groups at all the assessed post injection timepoints. At Day 14 and Day 90, decreases from baseline were greater for each active group compared to the corresponding placebo group. At Day 28, only the 500 µg/mL TenoMiR treatment group had a greater decrease from baseline compared to the corresponding placebo group. Despite showing greater decreases from baseline than placebo at most timepoints, there were no statistically significant differences in mean change from baseline in Quick-DASH Disability/Symptoms scores for all TenoMiR dose groups compared to placebo (Figures 2E-G). In the total study population, treatment with TenoMiR and placebo resulted in clinically relevant improvements in Quick-DASH Disability/Symptoms scores at Days 14,28 and 90 and TenoMiR treatment was significantly (p=0.05) better than placebo at Days 14 and 90 (Figure 2H).

Due to a lack of subject reporting in certain subdomains, mean ASES-E scores, changes from baseline could not be calculated and statistical analysis could not be performed for multiple parameters at numerous timepoints in the ASES-E. Where statistical analysis was possible, there were no significant differences in mean change from baseline in all ASES-E parameters for TenoMiR 500 µg/mL compared to the 500 µg/mL placebo group.

Compared to the corresponding placebo group, ASES-E scores were significantly higher for 200 µg/mL TenoMiR compared to the corresponding placebo group for supination in degrees - left at Day 90, extension left at Day 14, extension right at Day 14, flexion left at Day 14, flexion right at Day 14, grip strength left at Day 14, pronation left at Day 14, pronation right at Day 14, supination left at Day 14 and supination right at Day 14. Compared to the corresponding placebo group, ASES-E scores were significantly lower for 1500 µg/mL TenoMiR compared to the corresponding placebo group for extension in degrees – right at Day 14, pronation/supination arc – right at Day 14, Day 28, Day 90, and overall, and supination in degrees – left at Day 14, Day 28, Day 90, and overall.

Decreases from baseline in mean PRTEE Pain – Total Score were observed for all active treatment groups and all placebo groups at all the assessed post-injection timepoints. There was no apparent dose-related trend in the decreases from baseline for the active groups. For the 200 µg/mL TenoMiR group, the decreases from baseline were greater than those observed in the corresponding placebo group. For the 500 µg/mL and 1500 µg/mL TenoMiR groups the opposite was true, with larger decreases from baseline observed in the placebo groups. There were no statistically significant differences in mean change from baseline in PRTEE Pain – Total Score for all TenoMiR dose groups compared to placebo.

#### Imaging: UTC Ultrasound

All TenoMiR dosages resulted in improvements in echotypes I and II (approximately 15% at baseline to >60% 90 days post-treatment) at all timepoints post treatment a corresponding decrease in echotypes III and IV (approximately >80% at baseline to approximately 35% at 90 days post-treatment) was observed (Figures 3A-C).In contrast, placebo treated subjects showed mimimal t change in all echotypes from baseline (Figures 3A-C). Statistical analysis showed significant improvements in tendon structure compared to placebo at Day 28 (p<0.05) and Day 90 (p<0.01) reparative tendon (echotypes I and II) in TenoMiR treated subjects (Figure 3D). There was a corresponding significant reduction in degenerative tendon at Day 28 (p<0.05) and Day 90 (p<0.01) in TenoMiR treated subjects compared to placebo. There was a positive correlation (VAS vs % regenerative tendon on UTC) between decreasing pain and improved tendon structure for both the 250 µg/mL (p<0.05) and 500 µg/mL (p<0.001) while no correlation was seen at the 1500µg/mL dose (Figures 4 A-D). In the total study population, there was a significant (p<0.001) positive correlation between reduction in pain and improved tendon structure (Figure 4E). There was a positive correlation (QuickDASH vs % regenerative tendon on UTC) between improved upper limb function and improved tendon structure for both the 250 µg/mL (p<0.05) and 500 µg/mL (p<0.05) while no correlation was seen at the 1500µg/mL dose (Figures 4 F-I). In the total study population, there was a significant (p<0.001) positive correlation between improved upper limb function and improved tendon structure (Figure 4J).

**Figure 3.**
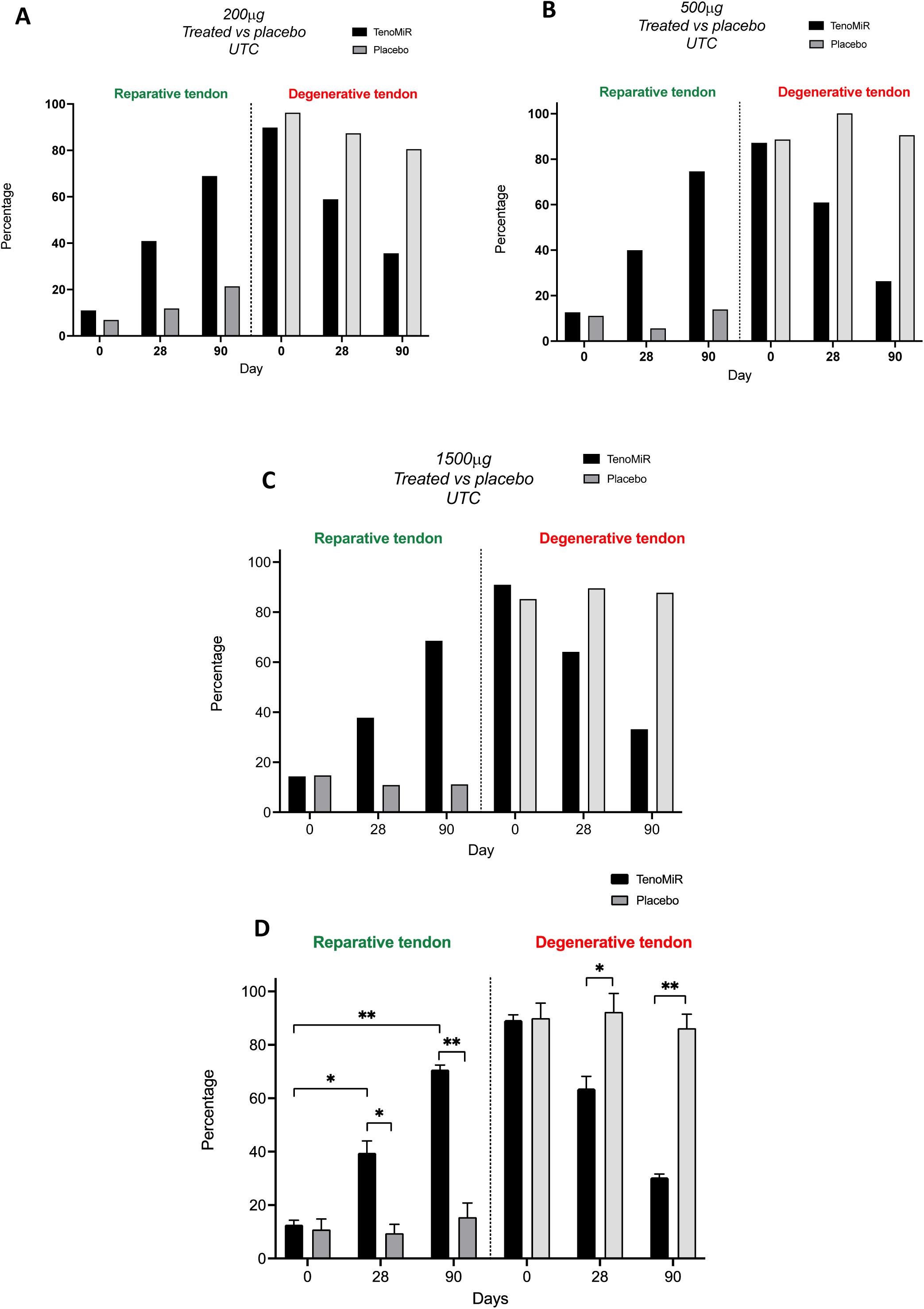
Ultrasound tissue Characterisation. Subanalysis (Type I and II echotypes, regenerative, Types III and IV echotypes degenerative) in (A-C) 200mcg,500mcg, 1500mcg (n=6 miR29a, n=2 placebo) and (D) overall patient population (n=18 miR29a, n=6 placebo)

**Figure 4.**
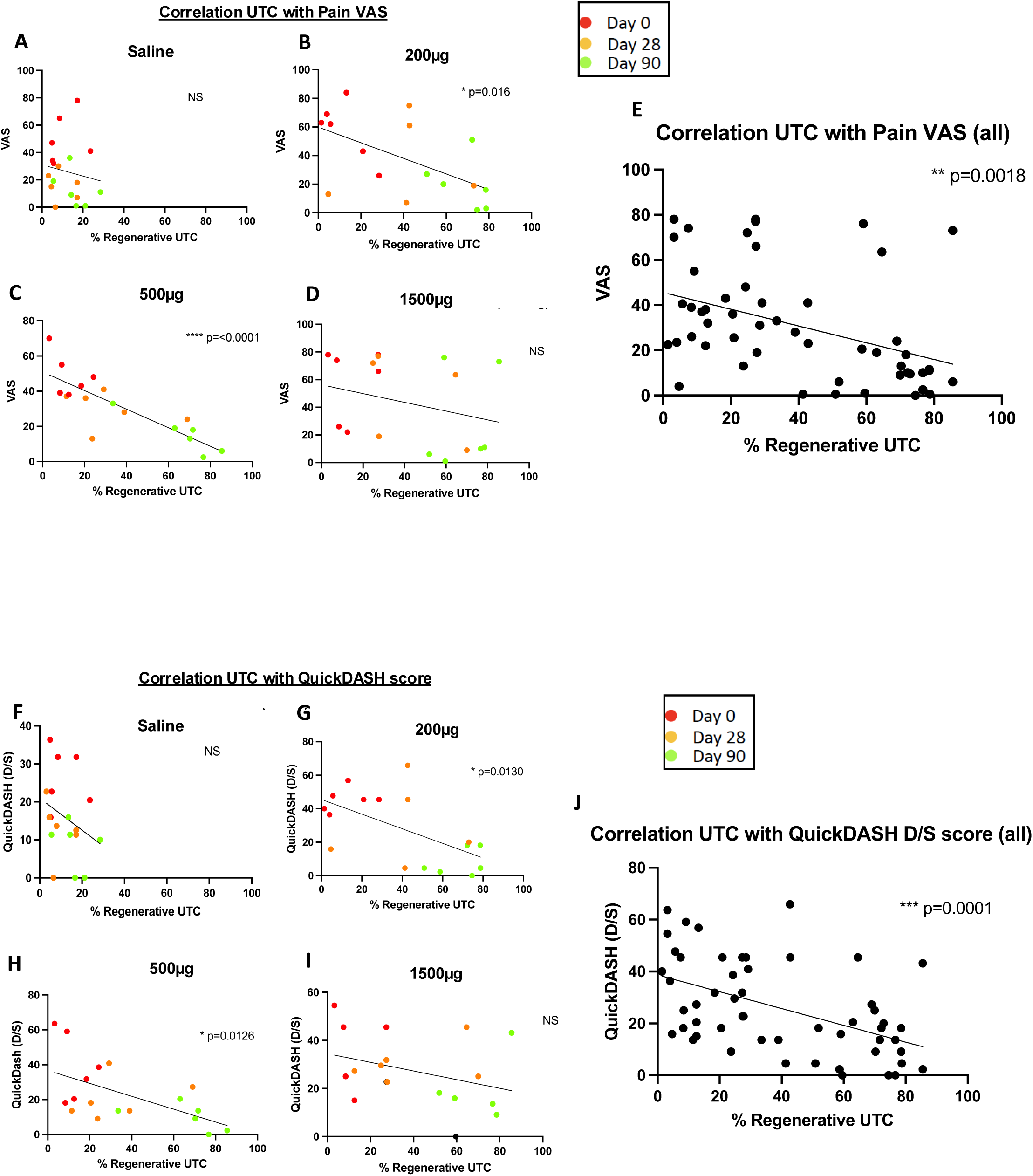
Correlation of QuickDASH with % of regenerative UTC. (A-D) Saline (Placebo), 200mcg,500mcg, 1500mcg and (E) overall patient population QuickDASH versus % of Echotype I and II on UTC (regenerative tendon)

## Discussion

In this study, one dose of 200, 500 or 1500 µg/mL of TenoMiR was well tolerated and no serious adverse events were noted in patients with lateral elbow tendinopathy. All adverse events were mild, and the majority was related to irritation around the skin of the injection site, which were transient and resolved within a few hours. Treatment with TenoMiR led to a substantial improvement in tendon structure at all doses over a 90 day period and there was improvement of symptoms and function in both groups with clear added benefit of TenoMiR treatment compared with placebo in the total study population in the QuickDASH upper limb functional score.

miRNAs are widely recognised as potent genetic regulators that influence diverse biological and developmental processes, while also holding a pivotal role in the pathogenesis of various diseases.(REF) This potency stems from a single miRNA’s ability to regulate entire cellular pathways by interacting with numerous target genes.^18^ Substantial improvements in designing, synthesis, binding affinity, stability, and target modulation effects of both miRNA mimics and anti-miRs have been accomplished through chemical changes to the nucleotide backbone since a major challenge for RNA-based therapeutic strategies is the possible degradation of oligonucleotides by RNases in extracellular and endocellular compartments. TenoMiR is a chemically synthesised mimic of miR29a where the sequence of the active (guide) strand of TenoMiR is identical to natural miR29a and therefore inhibits the expression of the same target genes. To increase stability and activity, TenoMiR’s backbone has been chemically modified via the introduction of 2′Fluro and 2′O-methyl groups and the passenger strand contains a 3′ cholesterol group to increase cellular uptake. TenoMiR has been shown to be rapidly degraded in human serum samples in an *in vitro* system (Gilchrist unpublished data). In a preclinical model, intravenous administration of TenoMiR showed negligible tissue distribution, even when administered at 25 times the level of the anticipated clinical dose. Following intra-tendinous injection in preclinical studies, similar negligible distribution in tissues is observed while systemically, levels of miR29a return to background levels after 6 hours. Additionally in patients with tendinopathy there was significantly reduced levels of miR29 in tendon biopsies and through mechanistic dissection in human in vitro studies miR29 was found to preferentially target collagen type 3 (Nat comm ref). Importantly, in this clinical study, the majority of adverse events were joint stiffness and pain in extremity in keeping with other musculoskeletal local injection therapies, and as such it was well tolerated in this group of patients.

The standard of care for tendinopathy in clinical practice remains conservative therapy based on minimizing exercise and anti-inflammatory medication. Novel methods have focused on regenerative medicine approaches with either platelet rich plasma (PRP) or stem cells.^19^ None of these recent ‘novel’ tendinopathy treatments have well-defined clinical rationale nor have they provided mechanistic insight; thus, their adoption within the clinical community has not been met with consensus. Herein, we show that local delivery of TenoMiR appears to mirror the concept of earlier lesion resolution and lack of lesion progression up to 3 months post treatment. This provides encouraging evidence that a miR manipulation can target inflammatory/matrix cross-talk in tendon disease and facilitate subsequent lesion resolution. The most significant challenge in treating tendinopathy, and indeed, many soft tissue pathologies is identifying a method to reset the pathological ‘switch’ that leads to irreversible degeneration. Functionally, miRs provide an added layer of complexity in tissue repair responses^20^ therefore directly manipulating these pathways allows precise targeting at the molecular level. In this instance we have utilized TenoMiR to directly modify collagen production via our previously published work on preferential targeting of collagen 3 and thereby recover the structural integrity of the tendon. Crucially, tendon degeneration occurs within a perpetuated inflammatory environment characterized by exaggerated cross-talk between immune cells and matrix producing stromal cells.^2^ Our data suggests that reintroducing TenoMiR into the tendon effectively mimics resetting the molecular ‘switch’ that regulates collagen production towards a reparative and homeostatic state. This, in turn, allows for a more effective healing response and improved tissue structure and function as evidenced by the positive correlation between improved upper limb function and tendon structure following TenoMiR treatment.

For the elbow pain/tendon pain questionnaire (VAS), Quick-DASH (DASH Disability/Symptom Scores and Calculated Work Scores) and PRTEE (all categories), decreases from baseline were observed for all active treatment groups, and corresponding placebo groups, at the majority of the assessed post-injection timepoints. The observed placebo effect resulted in isolated incidences of statistically significant differences in mean changes from baseline in multiple efficacy parameters. Additionally, there were no apparent TenoMiR dose-related trends in the decreases from baseline for the active groups for any efficacy parameter. This study offered an opportunity to probe further into the relationship between microRNA effector pathology and clinical presentation in and around the human tendon. Several observations suggest a plausible biological profile exhibited by miR29a in preclinical and translational studies of tendon tissues.^15^ Current literature surrounding the association between tendon structure, function and presentation of clinical signs, including pain, is ambiguous. Several meta-analyses demonstrate lack of consensus on whether rehabilitation protocols improve tendon structure as visualised by ultrasound.^21,22,23^ Recent studies indicate tendon abnormalities in asymptomatic tendons are predictive of future tendinopathy with a fourfold increased risk, this is in contrast to historical conjecture that tendon structure is unrelated to function.^24,25^ Successful restoration of function after tendon injury remains challenging clinically; as such, the ability of TenoMiR to directly modulate tendon structure as shown by the ultrasound data whilst improving functional scores is highly significant.

Importantly, randomised controlled trials (RCTs) in orthopaedics have evaluated the use of normal saline injections as a placebo/control group, expecting no physiological effect of this substance. However, as suggested by current best evidence within the field, questions arise as to whether normal saline can act as an appropriate placebo. ^18–21^ A recent systematic review of normal saline as placebo, specifically in lateral epicondylitis trials of similar design to this study, found that in all but one RCT examined there was no statistically significant different in PROMS between saline and non-saline (platelet-rich plasma [PRP], autologous conditioned plasma [ACP] corticosteroid and botulinum toxin) injection, with normal saline producing a clinically significant therapeutic effect.^22^ Based on this work it was proposed within the field that owing to the lack of robust evidence to elucidate the principles, physiological characteristics, effects and underlying mechanisms of normal saline injections, alternative control methods, such as sham syringes/needles, could be performed as better study control arms. Thus, interpretation of the efficacy results relating to PROMs in this study should be viewed with this caveat in mind, and our Phase 2 studies of TenoMiR will utilise sham injection (per regulatory scientific advice) to negate the significant placebo effect which makes clinical interpretation difficult in human tendon trials.

## Conclusion

In conclusion, TenoMiR administered in 200, 500 or 1500 µg/mL in a single intratendinous injection to patients with lateral elbow tendinopathy appeared safe, provided non dose-dependent plasma exposure of the drug, and resulted in significant improvement of tendon structure at 28 and 90 days post treatment. Despite presenting significant clinical and economic burden, tendinopathy remains notoriously difficult to treat due to the multifactorial nature of inciting factors. Tendon degeneration, resulting from injury and inflammation, has historically been considered irreversible and treatment strategies have focused on conservative management. Herein we demonstrate TenoMiR appears to facilitate lesion resolution in patients with lateral elbow tendinopathy up to three months post treatment which suggests potential disease modifying characteristics. This provides encouraging evidence that directly targeting inflammatory/matrix pathways can restore tendon structure and function, and highlights the potential of tendon modifying drugs promote early lesion resolution and superior tendon healing. Phase 2 studies are underway (NCT06192927) to establish the efficacy of TenoMiR in a larger population.

## Supporting information

Study outcome synopsis

Pharmcokinetics analysis plan

Statistical analysis plan

## Data Availability

All data produced in the present study are available upon reasonable request to the authors.

## Contributors

Study conception and design: NLM,IBM,ID,DB,DSG

Analysis and interpretation of the data: DB,PML,NLM,ID,

Drafting of the primary report: DB,PML, ID,DSG,NLM

Critical revision of the article for important intellectual content: NLM,DSG,ID,DB

Final approval of the article: all authors

Statistical expertise: MAC Clinical Research, NLM,DSG,ID

## Patient consent for publication

Not required.

## Ethics approval

The study was conducted according to the ethical principles of the Declaration of Helsinki and approved by the Wales REC1 (Health and Care Research Wales) Research Ethics Committee (REC 9/WA/0300) The study is registered with ClinicalTrials.gov (NCT04670289).

## Data availability statement

The datasets generated during and/or analysed at the end of the current study are not publicly available. Causeway is committed to sharing with qualified external researchers’ access to patient-level data and supporting clinical documents from eligible studies. These requests are reviewed and approved on the basis of scientific merit. All data provided are anonymised to respect the privacy of patients who have participated in the trial in line with applicable laws and regulations. The data may be requested from the corresponding author of the manuscript.

## References

1. Virta L, Joranger P, Brox JI, Eriksson R. Costs of shoulder pain and resource use in primary health care: a cost-of-illness study in Sweden. BMC Musculoskelet Disord 2012; 13: 17.

2. Millar NL, Murrell GAC, McInnes IB. Inflammatory mechanisms in tendinopathy - towards translation. Nat Rev Rheumatol 2017; 13(2): 110–22.

3. Boyd DF, Thomas PG. Towards integrating extracellular matrix and immunological pathways. Cytokine 2017; 98: 79–86.

4. Millar NL, Hueber AJ, Reilly JH, et al. Inflammation is present in early human tendinopathy. Am J Sports Med 2010; 38(10): 2085–91.

5. Millar NL, Dean BJ, Dakin SG. Inflammation and the continuum model: time to acknowledge the molecular era of tendinopathy. Br J Sports Med 2016.

6. Dakin SG, Martinez FO, Yapp C, et al. Inflammation activation and resolution in human tendon disease. Sci Transl Med 2015; 7(311): 311ra173.

7. Behzad H, Sharma A, Mousavizadeh R, Lu A, Scott A. Mast cells exert pro-inflammatory effects of relevance to the pathophyisology of tendinopathy. Arthritis Res Ther 2013; 15(6): R184.

8. John T, Lodka D, Kohl B, et al. Effect of pro-inflammatory and immunoregulatory cytokines on human tenocytes. J Orthop Res 2010; 28(8): 1071–7.

9. Dakin SG, Dudhia J, Smith RK. Science in brief: resolving tendon inflammation. A new perspective. Equine Vet J 2013; 45(4): 398–400.

10. Dakin SG, Dudhia J, Werling NJ, Werling D, Abayasekara DR, Smith RK. Inflamm-aging and arachadonic acid metabolite differences with stage of tendon disease. PLoS One 2012; 7(11): e48978.

11. Millar NL, Akbar M, Campbell AL, et al. IL-17A mediates inflammatory and tissue remodelling events in early human tendinopathy. Sci Rep 2016; 6: 27149.

12. He L, Hannon GJ. MicroRNAs: small RNAs with a big role in gene regulation. Nat Rev Genet 2004; 5(7): 522–31.

13. Bushati N, Cohen SM. microRNA functions. Annu Rev Cell Dev Biol 2007; 23: 175–205.

14. Bartel DP. MicroRNAs: target recognition and regulatory functions. Cell 2009; 136(2): 215–33.

15. Millar NL, Gilchrist DS, Akbar M, et al. MicroRNA29a regulates IL-33-mediated tissue remodelling in tendon disease. Nat Commun 2015; 6: 6774.

16. Bosch G, Rene van Weeren P, Barneveld A, van Schie HT. Computerised analysis of standardised ultrasonographic images to monitor the repair of surgically created core lesions in equine superficial digital flexor tendons following treatment with intratendinous platelet rich plasma or placebo. Vet J 2011; 187(1): 92–8.

17. Scott A, Docking S, Vicenzino B, et al. Sports and exercise-related tendinopathies: a review of selected topical issues by participants of the second International Scientific Tendinopathy Symposium (ISTS) Vancouver 2012. Br J Sports Med 2013; 47(9): 536–44.

18. Bushati N, Cohen SM. microRNA Functions. Annual Review of Cell and Developmental Biology 2007; 23(1): 175–205.

19. Miller LE, Parrish WR, Roides B, Bhattacharyya S. Efficacy of platelet-rich plasma injections for symptomatic tendinopathy: systematic review and meta-analysis of randomised injection-controlled trials. BMJ Open Sport & Exercise Medicine 2017; 3(1): e000237.

20. Rutnam ZJ, Wight TN, Yang BB. miRNAs regulate expression and function of extracellular matrix molecules. Matrix Biology 2013; 32(2): 74–85.

21. Murphy MC, Travers M, Chivers P, et al. Can we really say getting stronger makes your tendon feel better? No current evidence of a relationship between change in Achilles tendinopathy pain or disability and changes in Triceps Surae structure or function when completing rehabilitation: A systema. Journal of Science and Medicine in Sport 2023; 26(4-5): 253–60.

22. Murphy M, Travers M, Gibson W, et al. Rate of Improvement of Pain and Function in Mid-Portion Achilles Tendinopathy with Loading Protocols: A Systematic Review and Longitudinal Meta-Analysis. Sports Medicine 2018; 48(8): 1875–91.

23. Van Ark M, Rio E, Cook J, et al. Clinical Improvements Are Not Explained by Changes in Tendon Structure on Ultrasound Tissue Characterization After an Exercise Program for Patellar Tendinopathy. American Journal of Physical Medicine & Rehabilitation 2018; 97(10): 708–14.

24. McAuliffe S, McCreesh K, Culloty F, Purtill H, O’Sullivan K. Can ultrasound imaging predict the development of Achilles and patellar tendinopathy? A systematic review and meta-analysis. British Journal of Sports Medicine 2016; 50(24): 1516–23.

25. Docking SI, Cook J. Imaging and its role in tendinopathy: current evidence and the need for guidelines. Current Radiology Reports 2018; 6(11).

