## Supplementary material for "First in Human study of a microRNA29a mimic (TenoMiR) in patients with lateral elbow tendinopathy - a randomised, Placebo Controlled Phase 1 trial": Study outcome synopsis

### 2. SYNOPSIS

**Study Title:** A Phase 1, Single-Centre, Randomised, Double-Blind, Placebo-Controlled Study Evaluating the Safety, Tolerability and Pharmacokinetics of Single Ascending Doses of TenoMiR Injections in Subjects with Lateral Epicondylitis

**Sponsor:** Causeway Therapeutics

**Principal Investigator:** Dr Pui Man Leung MBChB MRCP(UK) FFPM DPM

**Study Site:** 1 study site located in the United Kingdom.

**Publication (reference):** Not applicable.

**Length of Study:**
Date of first subject entered: 14 August 2020
Date of last subject completed: 18 August 2021

**Phase:** 1

**Objectives:**
The primary objective of the study was to determine the safety and tolerability of single ascending doses of TenoMiR in subjects with lateral epicondylitis.

The secondary objectives of the study were:

- To determine the single dose pharmacokinetics (PK) of TenoMiR administration in subjects with lateral epicondylitis.
- To assess the efficacy of TenoMiR administration in subjects with lateral epicondylitis.

**Study Design:**
This was a Phase 1, randomised, double-blind, placebo-controlled study evaluating the safety, tolerability and PK of single ascending doses of TenoMiR injections in subjects with lateral epicondylitis. The study design is illustrated below:

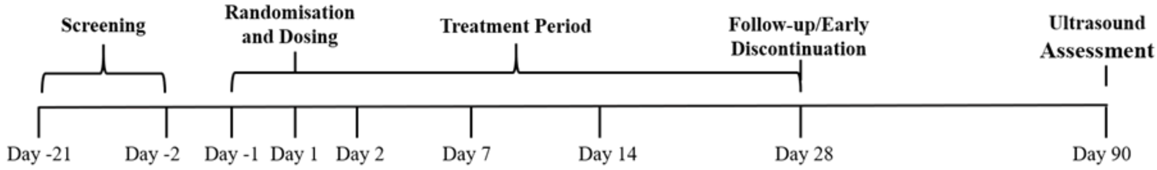


It was planned to enrol between 24 and 32 subjects (pending dose evaluation) with lateral epicondylitis. Subjects were divided into cohorts (8 subjects per cohort) and were randomised (3:1) to receive a single 1 mL dose of TenoMiR or saline placebo (0.9%) administered via injection into the lesion. The starting dose of TenoMiR was 200 µg/mL. It was planned to escalate the doses to 500 µg/mL and then 1500 µg/mL.

All doses were given as a 1 mL injection under ultrasound guidance into the affected area. In each cohort, no more than 2 subjects on the first dosing day (1 active; 1 placebo) were dosed, such that no more than 1 subject received an active TenoMiR dose for the first time at each dose level. After 72 hours, depending on the safety and tolerability of the previously dosed sentinel subjects, dosing continued in the remaining subjects (6 subjects; 5 active, 1 placebo) at the same dose level in each cohort.

Doses were administered in an escalating manner, following satisfactory review of all safety, tolerability and plasma PK data from lower doses. Except for the starting dose, the dose levels studied were preliminary, with actual subsequent doses determined based on ongoing evaluations of the safety, tolerability, plasma PK and efficacy data (where available) by the Safety Review Committee.

Subjects were required to attend the Clinical Research Unit (CRU) up to 28 days prior to Day 1 for a Screening visit to ensure they met the inclusion/exclusion criteria and were otherwise in good health. The ultrasound assessment at Screening was conducted at any time during the 4-week Screening period and included a scan of the contra-lateral elbow. At least 1 week prior to dosing, subjects discontinued any use of opiate or non-steroidal anti-inflammatory drug (NSAID) medications they were using for pain management. From Day -7 to Day -1, subjects were allowed to take paracetamol (less than or equal to 4 g/day) or use an ice pack to manage pain.

Subjects attended the CRU on Day -2/Day -1 and predose on Day 1 to confirm eligibility and undergo safety and efficacy baseline assessments. Following confirmation of eligibility, subjects were randomised and dosed with TenoMiR or placebo. After the injection, subjects were rested for 30 minutes and were advised against massage or hot fomentation. Ice packs and paracetamol (less than or equal to 4 g/day) may have been used in case of any immediate and persistent discomfort. Subjects underwent postdose safety and PK assessments and were discharged from the CRU on the same day. Subjects were allowed to remain at the CRU overnight at the discretion of the Investigator on Day 1, if required for safety reasons.

Subjects returned to the CRU for PK sampling assessments on Day 2, safety and PK assessments on Day 7, and safety and efficacy assessments were performed on Day 14, Day 28 and Day 90. Subjects also underwent ultrasound assessments on Day 28 and Day 90. To mitigate the risks to subject safety from severe acute respiratory syndrome coronavirus 2 (SARS-CoV-2), if required and where possible, Follow-up visits were conducted remotely by CRU staff.

The duration of the study for each subject was approximately 16 weeks.

**Number of Subjects:**

**Planned:** 24 to 32
**Randomised:** 24
**Treated:** 24
**Completed:** 24

**Diagnosis and Main Criteria for Inclusion:**
Male or female subjects aged between 18 and 70 years (inclusive, with no evidence of skeletal immaturity) and a body mass index of 18 to 35 kg/m$^2$ (inclusive). Subjects must have had a clinical diagnosis of lateral epicondylitis, with symptoms that could be reproduced with resisted supination or wrist dorsiflexion (as confirmed by tenderness at lateral epicondyle and positive pick up back of chair sign). In addition, subjects' symptoms must have persisted for at least 6 weeks to 6 months despite conservative treatment that included one or combinations of physical therapy, splinting and/or NSAIDs.

**Study Drug, Dose and Mode of Administration:**
The Investigational Medicinal Product (IMP) was provided as 2 mL clear glass Type I injection vials containing 1.5 mL of clear IMP/drug solution at a concentration of 5 mg/mL. Each vial was stoppered with a 13 mm Flurotec injection stopper with a 13 mm aluminium overseal flip-off cap. The labelling was Annex 13 compliant.

The IMP was administered once on Day 1 using ultrasound guided injections (volume = 1 mL) into the common extensor tendon of the affected elbow, by site staff experienced in this technique.

Ultrasonography was applied using a 5-7.5 MHz linear array transducer on a LOGIQ E9 Ultrasound System. The probe was placed along the lateral aspect of the elbow parallel to the longitudinal axis of the common extensor tendon. The radial head, radiohumeral joint, lateral epicondyle and common extensor tendon were visualised. Aseptic conditions around the injection site were maintained using a sterile jelly applied to the skin and the ultrasound probe was enclosed in a sterile cover. After visualisation of the tip of the needle at the exact site (hypointense area), 1 mL of TenoMiR was injected.

To help with any discomfort from the injection, a subcutaneous dose of up to 5 mL of 1% lignocaine (without adrenaline) could have been administered at the discretion of the Investigator. After the injection, the subject was rested for 30 minutes and was advised against massage or hot fomentation. Ice packs and paracetamol (less than or equal to 4 g/day) could have been used in case of any immediate and persistent discomfort.

**Reference Therapy, Dose and Mode of Administration:**
The reference product was saline placebo (0.9%).

Placebo was administered once on Day 1 using ultrasound guided injections (volume = 1 mL) into the common extensor tendon of the affected elbow, by site staff experienced in this technique.

Ultrasonography was applied using a 5-7.5 MHz linear array transducer on a LOGIQ E9 Ultrasound System. The probe was placed along the lateral aspect of the elbow parallel to the longitudinal axis of the common extensor tendon. The radial head, radiohumeral joint, lateral epicondyle and common extensor tendon were visualised. Aseptic conditions around the injection site were maintained using a sterile jelly applied to the skin and the ultrasound probe was enclosed in a sterile cover. After visualisation of the tip of the needle at the exact site (hypointense area), 1 mL of placebo was injected.

To help with any discomfort from the injection, a subcutaneous dose of up to 5 mL of 1% lignocaine (without adrenaline) could have been administered at the discretion of the Investigator. After the injection, the subject was rested for 30 minutes and was advised against massage or hot fomentation. Ice packs and paracetamol (less than or equal to 4 g/day) could have been used in case of any immediate and persistent discomfort.

**Duration of Treatment:** 1 day (Day 1)

**Criteria for Evaluation:**
The primary endpoint of the study was the comparison of safety data between TenoMiR versus placebo as measured by incidence of adverse events (AEs), clinical laboratory abnormalities, changes in vital signs (blood pressure, temperature, respiratory rate and pulse rate), 12-lead electrocardiogram (ECG) parameters, physical examinations and skin score assessment at 14 days post-injection.

The secondary endpoints of the study were:

- The plasma PK of TenoMiR as shown by maximum drug plasma concentration ($C_{max}$), time to reach $C_{max}$ ($t_{max}$) and area under the plasma vs. concentration time curve (AUC).
- Efficacy of a single dose of TenoMiR on elbow pain as measured using Visual Analogue Scale (VAS) from predose on Day 1 to Day 14, Day 28 and Day 90.
- Efficacy of a single dose of TenoMiR on disability and symptoms measured using the Disabilities of the Arm, Shoulder and Hand (Quick DASH) Score from predose on Day 1 to Day 14, Day 28 and Day 90.
- Efficacy of a single dose of TenoMiR on pain and disability as measured by the American Shoulder and Elbow Surgeons Elbow (ASES-E) Score from predose on Day 1 to Day 14, Day 28 and Day 90.
- Efficacy of a single dose of TenoMiR on pain and disability as measured by the Patient Rated Tennis Elbow Evaluation (PRTEE) from predose on Day 1 to Day 14, Day 28 and Day 90.
- Efficacy of a single dose of TenoMiR on lateral elbow tendons as measured by change from baseline ultrasound assessment from predose on Day 1 to Day 28 and Day 90.

**Evaluation Methods:**
Safety was assessed through AE reporting, 12-lead ECG, skin score assessments, vital signs, physical examinations and clinical laboratory evaluations.

The PK of TenoMiR was assessed through serial blood samples to determine plasma concentration of miR29a.

Efficacy was assessed serially by VAS pain reporting, ASES-E score for the assessment of shoulder and elbow function, Quick DASH score, PRTEE score, and ultrasound assessment of the lateral elbow tendon.

**Statistical Methods:**
Safety parameters were listed and summarised using descriptive statistics.

Pharmacokinetic data were listed for each subject, along with summary statistics including arithmetic and geometric means, standard deviations, minimum, maximum and median values, and coefficients of variation.

Efficacy parameters were listed and summarised using descriptive statistics. Effects of TenoMiR on efficacy parameters were analysed with a generalised linear mixed model (GLMM) using an efficacy parameter as responsible variable, treatment (active and placebo), timepoint, interaction between treatment and timepoint as mixed effects, baseline measurements as a covariate, and subject and elbow (left and right) as random effects. This model was applied to each efficacy parameter. Treatment difference between active and placebo at different timepoints together with 95% confidence intervals (CIs) was derived from the GLMM. For a continuous outcome, treatment difference was measured as the difference in least squares mean; for a binary outcome, treatment difference was measured as the odds ratio.

**Summary:**

**<u>Results</u>**

**<u>Study Population</u>**
Twenty-four subjects (16 male subjects and 8 female subjects) were enrolled, randomised evenly across three dose cohorts and dosed during the study. All 24 subjects completed the study.

**Safety Results**

Single injections of TenoMiR appeared to be well tolerated over the 200 to 1500 µg/mL dose range, and there were no safety concerns that could be attributed to treatment with TenoMiR. No deaths were reported during the study and one serious AE (SAE) was reported, a suspected SARS-CoV-2 infection that required hospitalisation in the 200 µg/mL TenoMiR treatment group. No subjects discontinued due to a treatment-emergent AE (TEAE).

Overall, 18 (75.0%) subjects experienced 59 TEAEs across all treatment groups. Of the 59 events reported, 48 events reported by 18 (75.0%) subjects were mild in severity, 7 events reported by 5 (20.8%) subjects were moderate and 4 events reported by 1 (4.2%) subject were severe. All 4 severe events were reported by 1 (4.2%) subject in the 200 µg/mL TenoMiR treatment group and were not considered to be related to treatment.

The most commonly reported TEAEs were within the musculoskeletal and connective tissue disorders and nervous system disorders System Organ Classes (SOCs). Within these SOCs, the most common TEAEs (by preferred term [PT]) were:

- Headache (8 [33.3%] subjects; 2 [33.3%] subjects each in the 200 µg/mL and 1500 µg/mL TenoMiR treatment groups, 3 [50.0%] subjects in the 500 µg/mL TenoMiR treatment group, and 1 [50.0%] subject in the 1500 µg/mL placebo group).
- Arthralgia (6 [25.0%] subjects; 1 [16.7%] subject in the 200 µg/mL TenoMiR treatment group, 3 [50%] subjects in the 500 µg/mL TenoMiR treatment group, and 1 [50%] subject each in the 200 µg/mL and 500 µg/mL placebo groups).
- Joint stiffness (2 [8.3%] subjects; 2 [33.3%] subjects in the 500 µg/mL treatment group).
- Pain in extremity (2 [8.3% subjects]; 1 [16.7%] subject in the 200 µg/mL TenoMiR treatment group and 1 [50.0%] subject in the 200 µg/mL placebo group).

With the exception of contusion (2 [8.3%] subjects; 1 [16.7%] subject each in the 200 µg/mL TenoMiR and 500 µg/mL TenoMiR treatment groups), all other TEAEs were reported by a single subject.

Overall, 8 of the 59 reported TEAEs were considered to be possibly or probably related to treatment. Five of the 8 treatment related TEAEs were reported by subjects administered TenoMiR, and 3 were reported by subjects administered placebo. There were no treatment-related TEAEs reported at the highest dose level (1500 µg/mL TenoMiR) or in the 1500 µg/mL placebo group.

Treatment-related TEAEs were reported within the general disorders and administration site conditions, injury, poisoning and procedural complications, and musculoskeletal and connective tissue disorders SOCs. Within these SOCs, the only treatment-related TEAEs (by PT) reported by more than 1 subject were:

- Arthralgia (3 [12.5%] subjects; 1 [16.7%] subject reported 1 event in the 500 µg/mL TenoMiR treatment group, 1 [50.0%] subject reported 2 events in the 200 µg/mL placebo group and 1 [50.0%] subject reported 1 event in the 500 µg/mL placebo group).
- Joint stiffness (2 [8.3%] subjects; 2 [33.3%] subjects reported 2 events in the 500 µg/mL TenoMiR treatment group).

Single treatment-related events of injection site joint pain and epicondylitis were reported in the 500 µg/mL TenoMiR treatment group and the 200 µg/mL TenoMiR treatment group, respectively.

There was a significantly greater incidence of TEAEs within the nervous system disorders SOC reported by subjects administered TenoMiR compared with subjects administered placebo. Overall, 10 (41.7%) subjects reported 15 nervous system disorder TEAEs. Of the 15 events, 14 were reported by 9 subjects administered TenoMiR (2 [33.3%] subjects administered 200 µg/mL TenoMiR reported 3 events, 5 subjects administered 500 µg/mL TenoMiR reported 9 events, and 2 [33.3%] subjects administered 1500 µg/mL TenoMiR reported 2 events). There was only 1 nervous system disorder TEAE reported by a subject administered placebo (1 [50.0%] subject reported 1 event in the 1500 µg/mL placebo group).

The only TEAEs (by PT) that were reported by more than one subject administered TenoMiR were headache, arthralgia, contusion and joint stiffness. Eleven events of headache were reported by 8 (33.3%) subjects, 7 of whom were administered TenoMiR (2 [33.3%] subjects reported 3 events in the 200 µg/mL TenoMiR treatment group, 3 [50.0%] subjects reported 5 events in the 500 µg/mL TenoMiR treatment group, and 2 [33.3%] subjects reported 2 events in the 1500 µg/mL TenoMiR treatment group). Both events of joint stiffness were reported in the 200 µg/mL TenoMiR treatment group, and both were considered to be related to treatment.

There were no significant treatment- or dose-related trends in clinical laboratory evaluations, vital signs or ECG parameters. Four clinically significant findings in the physical examinations performed were reported by 2 subjects (200 µg/mL TenoMiR treatment group and 200 µg/mL placebo group), however none were considered to be related to treatment.

There was a significantly greater number of skin score assessments above Grade 0 reported by subjects administered 500 µg/mL TenoMiR compared with all other treatment groups. The greatest incidence of an increased score was at 1 hour postdose, where 6 (25.0%) subjects reported a Grade 1 score for pain. The highest score overall was a Grade 3 score for tenderness, reported by 1 subject administered 500 µg/mL TenoMiR.

**Pharmacokinetic Results**

Following a single injection of TenoMiR at 200 µg/mL, 500 µg/mL pr 1500 µg/mL, all subjects had a quantifiable value of miR29a in plasma up to 144 hours postdose.

Following a single injection of TenoMiR, geometric mean $C_{max}$ values of miR29a of 3.16, 6.63 and 2.37 were observed following administration of 200, 500 and 1500 µg/mL TenoMiR, respectively. Geometric mean $C_{max}$ values were attained on average (median $t_{max}$) at 13.5, 1.26 and 0.75 hours postdose following administration of 200, 500 and 1500 µg/mL TenoMiR, respectively. Following a single injection of placebo, a geometric mean $C_{max}$ value of miR29a of 1.90 was observed and was attained on average (median $t_{max}$) at 7 hours postdose.

Systemic exposure to miR29a ($C_{max}$ and AUC from time zero to the last quantifiable concentration [$AUC_{0-t}$]) tended to decrease with dose; however, there was no apparent dose proportional relationship. Minimum observed drug plasma concentration ($C_{min}$) values decreased with dose and the change was less than dose proportional. For a doubling in dose, $C_{min}$ was predicted to decrease 1.6-fold and the 90% CIs [1.22, 2.07]) were outside the prescribed limits of 1.6 to 2.5; therefore, dose proportionality could not be confirmed.

The relationship between $C_{max}$, $AUC_{0-t}$ and $C_{min}$ values and dose following a single injection of TenoMiR to subjects with lateral epicondylitis are presented below.

| Dose (µg/mL) | Fold Difference in Dose | $C_{max}$ | Fold Decrease | $AUC_{0-t}$ | Fold Decrease | $C_{min}$ | Fold Decrease |
|---|---|---|---|---|---|---|---|
| 200 | 1 | 3.16 | 1 | 286 | 1 | 0.62 | 1 |
| 500 | 2.5 | 6.63 | 0.48 | 229 | 1.25 | 0.27 | 2.28 |
| 1500 | 3 | 2.37 | 2.80 | 172 | 1.33 | 0.16 | 1.70 |
| **Overall** | 7.5 | | 1.33 | | 1.66 | | 3.88 |

Abbreviations: $AUC_{0-t}$ – area under the values versus time curve from time zero to the last quantifiable value; $C_{max}$ – maximum observed values; $C_{min}$ – minimum observed values.

Estimates of the exponent of the power model (and 90% CIs) are presented below.

| PK Parameter | Exponents[a] | | For a Doubling in Dose | | LOF |
|---|---|---|---|---|---|
| | Estimate | 90% CI | Estimate | 90% CI | p-value |
| $C_{min}$ | -0.666 | -1.05, -0.281 | 1.59 | 2.07, 1.22 | 0.183 |

Abbreviations: CI – confidence intervals; $C_{min}$ – minimum observed values; LOF – lack of fit (p-value <0.05 would indicate lack of fit).
a Exponent from the power model.

Between-subject variability in the extent of systemic exposure to miR29a was considered to be high following administration of 200 and 500 µg/mL TenoMiR; geometric coefficients of variation (CVs) of 99.9% and 154% for $C_{max}$, and 119% and 111% for $AUC_{0-t}$, were observed following administration of 200 and 500 µg/mL TenoMiR, respectively. Between-subject variability in the extent of systemic exposure to miR29a was considered to be moderate following administration of 1500 µg/mL TenoMiR; geometric CVs of 53.1% for $C_{max}$, and 31.5% for $AUC_{0-t}$, were observed. Between-subject variability in the extent of systemic exposure to miR29a was considered to be high following administration of placebo, with geometric CVs of 91.5% and 66.5% for $C_{max}$ and $AUC_{0-t}$, respectively.

**Efficacy Results**

For the majority of efficacy parameters, decreases from baseline were observed for all active treatment groups and all placebo groups at all the assessed post-injection timepoints. There were no apparent dose-related trends in the decreases from baseline for the active groups for any efficacy parameter.

For the elbow pain/tendon pain questionnaire (VAS), Quick DASH (DASH Disability/Symptom Scores and Calculated Work Scores) and PRTEE (all categories), decreases from baseline were observed for all active treatment groups, and corresponding placebo groups, at the majority of the assessed post-injection timepoints and no dose-related trends were observed. The observed placebo effect resulted in isolated incidences of statistically significant differences in mean changes from baseline in multiple efficacy parameters. Additionally, there were no apparent TenoMiR dose-related trends in the decreases from baseline for the active groups for any efficacy parameter.

A placebo effect was observed for the elbow pain/tendon pain questionnaire (VAS). At the majority of timepoints, the placebo groups had larger decreases from baseline compared to the corresponding TenoMiR dose groups; at all other timepoints, changes from baseline were comparable between TenoMiR dose groups and the corresponding placebo groups. However, TenoMiR treatment at all dosages and timepoints resulted in a Minimally Clinically Important Difference (MCID) of greater than 11 points change in the VAS score from baseline. Due to sample size in this Phase 1b study, no instances of statistically significant differences in change from baseline were observed for TenoMiR dose groups compared to placebo.

For all PRTEE parameters, the decrease from baseline was greater in the 200 µg/mL TenoMiR dose group than those observed in the corresponding placebo group; for the 500 µg/mL and 1500 µg/mL TenoMiR dose groups the opposite was true, with larger decreases from baseline observed in the placebo groups. Additionally, there were no instances of statistically significant differences in mean change from baseline for all TenoMiR dose groups compared to corresponding placebo groups. However, the TenoMiR treatment at all dosages and timepoints resulted in a MCID of greater than 11 points change in the PRTEE score from baseline.

No clear dose-related trends were observed in the mean change from baseline scores in any Quick DASH category. Despite greater decreases from baseline for each active group compared to the corresponding placebo groups on Day 14 and Day 90, there were no significant differences in mean change from baseline in DASH Disability/Symptom Scores for all TenoMiR dose groups compared to placebo, due to a placebo effect. TenoMiR treatment at all dosages and timepoints resulted in a MCID of greater than 20 points change in the Quick DASH score from baseline. For Calculated Work Scores, there was no discernible effect of TenoMiR; a statistically significant difference in the mean change from baseline for TenoMiR 500 µg/mL on Day 14 compared to placebo was observed, however this was an isolated result. For the DASH Sports/Performing Arts Module Scores, decreases from baseline were observed for the 200 µg/mL and 500 µg/mL TenoMiR treatment groups; however, the 1500 µg/mL TenoMiR dose group had statistically significant increases from baseline compared to placebo on Day 14 and Day 28, and on average.

Where statistical analysis was possible for ASES-E parameter scores, there were no apparent dose-related trends observed. Statistically significant differences in ASES-E parameters for the 200 µg/mL and 1500 µg/mL TenoMiR treatment groups were sporadic, and there were no significant differences in mean change from baseline for TenoMiR 500 µg/mL compared to the 500 µg/mL placebo group. Due to a lack of subject reporting in certain subdomains, mean ASES-E scores, changes from baseline could not be calculated and statistical analysis could not be performed for multiple ASES-E parameters at numerous timepoints.

Recently, a novel imaging modality called Ultrasound Tissue Characterisation (UTC) has been developed to visualise tendon structure and to quantify tendon matrix integrity. Unlike two-dimensional ultrasound and colour Doppler, UTC objectively quantified grey-scale tendon matrix changes into 4 different echotypes related to tendon integrity. As illustrated in the figure below, Types I (green) and II (blue) represent an organised matrix; Types III (red) and IV (black) represent a disorganised matrix.

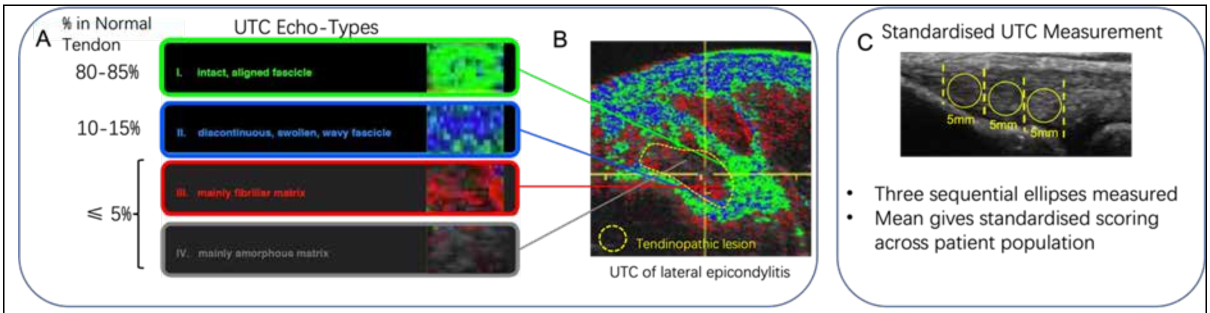


All TenoMiR dosages resulted in improvements in echotypes I and II (approximately 15% at baseline to >60% 90 days post-treatment) at all timepoints post-treatment whilst also causing a decrease in echotypes III and IV (approximately >80% at baseline to approximately 35% at 90 days post-treatment). No clear dose-related trend was observed. Placebo (saline treatment) resulted in little change in echotype from baseline and in particular caused little change in echotypes I and II. Statistical analysis shows significant improvements in tendon structure at Day 28 ($p<0.05$) and Day 90 ($p<0.01$) of reparative tendon (echotypes I and II) in TenoMiR treated subjects compared to placebo. There was a corresponding significant reduction in degenerative tendon at Day 28 ($p<0.05$) and Day 90 ($p<0.01$) in TenoMiR treated subjects compared to placebo.

**Conclusions**

- Single injections of TenoMiR appeared to be safe and well tolerated when administered over a dose range of 200 to 1500 µg/mL, with no dose-related trend observed in the frequency of TEAEs.
- No deaths or TEAEs leading to withdrawal occurred in the study. One SAE of a suspected SARS-CoV-2 infection that required hospitalisation was reported by 1 subject in the 200 µg/mL TenoMiR treatment group. The event was not considered to be related to treatment.
- Overall, 18 (75.0%) subjects experienced 59 TEAEs, 8 of which were considered to be either possibly or probably related to treatment. The majority of events were mild in severity. Four severe events were reported by 1 subject in the 200 µg/mL TenoMiR treatment group, however they were not considered to be related to treatment.
- There were no significant treatment- or dose-related trends in clinical laboratory evaluations, vital signs or ECG parameters. Four clinically significant physical examination findings were reported by 2 subjects but were not considered to be related to treatment.
- Between-subject variability in the extent of systemic exposure to miR29a was considered to be moderate to high.
- Geometric mean $C_{max}$ and $AUC_{0-t}$ values tended to decrease with increasing dose; however, there was no apparent dose proportional relationship.
- $C_{min}$ values decreased with dose and $C_{min}$ was predicted to decrease 1.6-fold; the 90% CIs were outside the prescribed limits of 1.6 to 2.5.
- For the majority of efficacy parameters, decreases from baseline were observed for all active treatment groups and all placebo groups at all the assessed post-injection timepoints.
- At all timepoints, TenoMiR treatment irrespective of dose resulted in a MCID in all subject reported outcomes (VAS, PRTEE, Quick DASH).
- There were no apparent TenoMiR dose-related trends in the decreases from baseline for the active groups for any efficacy parameter.
- There were no statistically significant differences in mean change from baseline for the elbow pain/tendon pain questionnaire VAS scores for all TenoMiR dose groups compared to placebo, due to the placebo effect.
- No clear dose-related trends were observed in the mean change from baseline scores for Quick DASH categories.
- There were no statistically significant differences in mean change from baseline in all PRTEE parameters for all TenoMiR dose groups compared to placebo.
- Statistically significant improvements in tendon structure at Days 28 and 90 post-treatment with all dosages of TenoMiR.

**Date of the Report:** 31 March 2022
