## Supplementary material for "First in Human study of a microRNA29a mimic (TenoMiR) in patients with lateral elbow tendinopathy - a randomised, Placebo Controlled Phase 1 trial": Pharmcokinetics analysis plan

### PHARMACOKINETIC ANALYSIS PLAN

#### A Phase 1, Single-Centre, Randomised, Double-Blind, Placebo-Controlled Study Evaluating the Safety, Tolerability and Pharmacokinetics of Single Ascending Doses of TenoMiR Injections in Subjects with Lateral Epicondylitis Investigational Medicinal Product: TenoMiR

| | |
|---|---|
| **KinetAssist Reference Number:** | MPC101 |
| **Version** | 1.0 |
| **Clinical Protocol Number:** | CWT-TE1 |
| **MAC Number:** | MAC 075 |
| **Test Article** | TenoMiR |
| **Responsible Scientist:** | Charlie Brindley, PhD<br>KinetAssist Limited<br>Larchwood, Shieldhill Road<br>Quothquan,<br>Lanarkshire. ML12 6NA, UK |
| **Sponsor:** | Causeway Therapeutics<br>Imaging Centre of Excellence Building<br>Queen Elizabeth University Hospital<br>Glasgow, G51 4TF, UK |
| **Proposed Analysis Start Date:** | 2020 |
| **Proposed Analysis Completion Date:** | 2020 |
| **Pharmacokinetic Report Appendix:** | 2020 |

**Pharmacokinetic Plan Approval**

**On behalf of KinetAssist Limited**

_______________________ _______________

Charlie Brindley
Responsible Scientist
Director, KinetAssist Limited

Date

**On behalf of MAC Clinical Research Manchester (Early Phase Unit)**

_______________________ _______________

Paul Steven
Statistician

Date

**On behalf of Causeway Therapeutics**

Derek Gilchrist 2 SEPT 2020

[Name] DEREK GILCHRIST
[Title] CEO

Date

### 1 CLINICAL STUDY OBJECTIVES

#### Primary Objectives

The primary objective of this study is to determine the safety and tolerability of single ascending doses of TenoMiR in subjects with lateral epicondylitis.

#### Secondary Objectives

The secondary objectives of this study are:

- To determine the single dose pharmacokinetics (PK) of TenoMiR in subjects with lateral epicondylitis.
- To explore the efficacy of a single dose of TenoMiR in subjects with lateral epicondylitis.

This PK Analysis Plan (PAP) describes the PK analysis and reporting of plasma concentrations of miR29a (measured as relative expression of miR29a compared to miR159a control following single injections of TenoMiR).

### 2 CLINICAL STUDY DESIGN

This is a phase 1, randomised, double-blind, placebo-controlled study evaluating the safety, tolerability and PK of single ascending doses of TenoMiR injections in subjects (male or female participants aged between 18 and 70 years) with lateral epicondylitis. It is planned to enrol between 24 and 32 subjects (pending dose evaluation) with lateral epicondylitis. Subjects will be divided into cohorts (8 subjects per cohort) and will be randomised (3:1) to receive a single 1 mL dose of TenoMiR or saline placebo (0.9%) administered via injection. The starting dose of TenoMiR will be 200 µg/mL. The target doses for escalation will be 500 µg/mL and then 2000 µg/mL. One additional cohort (8 subjects) may be utilised to explore an additional dose, if necessary. The decision to explore an additional dose will be made by an interim analysis group comprising the Sponsor and Safety Review Committee (SRC). Except for the starting dose, the doses outlined are preliminary, with actual subsequent doses determined based on the ongoing evaluation of the safety, tolerability, plasma PK and efficacy data (where available) by the SRC. Sentinel dosing (1 active; 1 placebo) will occur at every dose level. After 72 hours, depending on the safety and tolerability of the previously dosed sentinel subjects, dosing will continue in the remaining subjects (6 subjects; 5 active, 1 placebo) at the same dose level in each cohort.

#### Blood Sampling for Pharmacokinetic Analysis

Blood samples will be taken from each subject at pre-dose and at 0.5, 1, 2, 3, 4, 8, 12, 24 and 36 h post-dose on Day 1. An additional sample will be taken 7 days (± 1 day) after dosing on Day 1. Plasma concentrations of miR29a (i.e. delta Ct values) will be quantified using a validated quantitative real time PCR method developed at Thermo Fisher.

### 3 GLP AND GCP COMPLIANCE

#### 3.1 Compliance statement (Statement of Authenticity)

"This pharmacokinetic analysis was conducted in a facility which operates a Quality Management System (QMS). The QMS has been designed to be compatible with Good Clinical Practice (GCP) requirements. This work has been subject to an independent Quality Assurance audit."

#### 3.2 Amendments to and deviations from the Pharmacokinetic Analysis Plan (PAP)

Amendments to the agreed PAP, whether instigated by KinetAssist or by the Sponsor, will be documented and sent to the Sponsor for authorisation. Any deviations from the analysis populations or analysis methodology detailed in the PAP will be documented in the raw data and final report appendix.

#### 3.3 Quality assurance

The work described in this analysis plan, including the Report Appendix, will be subject to Quality Assurance evaluation by Tower Mains Limited, Edinburgh, UK. Data analysis and reporting will undergo an independent Quality Control (QC) review under the supervision of KinetAssist Limited, according to KinetAssist's SOP-08 [1], prior to provision of the Draft PK Report Appendix. The QC review will be carried out by Quanticate Limited, Edinburgh, UK.

### 4 ENDPOINTS

#### 4.1 Pharmacokinetic endpoints

The primary endpoints will be $C_{max}$ and $AUC_{0\text{-}t}$.

#### 4.2 Other endpoints

Other endpoints are listed in Section 6.2.

### 5 ANALYSIS POPULATION

The PK Population is defined as all subjects who received a dose of TenoMiR on Day 1 and have evaluable plasma concentrations of miR29a.

PK parameters will be excluded from the analysis and summary statistics, where there are insufficient plasma concentration data available.

### 6 PHARMACOKINETIC ANALYSIS

#### 6.1 Overall considerations

Plasma concentration data will be provided by the Sponsor in Excel format as delta Ct values. Relative expression of miR29a will be calculated as follows:

| Endpoint Name | Derivation | Description |
|---|---|---|
| Relative expression of miR29a compared to miR159a Control | $= 2^{-\Delta Ct}$<br><br>where ΔCt is defined as;<br><br>ΔCt (delta Ct) = miR29a Ct – miR159a Ct. | Calculated for each subject for each timepoint, including baseline.<br><br>Represents relative expression of miR29a against miR159a. |

Blood sampling times, demographic data and randomisation schedules will be provided by the Sponsor.

#### 6.2 Pharmacokinetic parameter estimation

Definition and estimation of PK parameters are described in KinetAssist's SOPs [2,3]. PK parameters will be estimated for each subject using a fully validated version of WinNonlin Phoenix (Version 8.3) [4] (or higher version). The following parameters will be derived, where appropriate, from the individual plasma concentration (i.e. relative expression values) versus time profiles of miR29a.

| Parameter | Definition |
|---|---|
| $C_{max}$ | The maximum observed concentration. |
| $C_{min}$ | The minimum observed concentration. |

| | |
|---|---|
| $C_{ave}$ | The average of plasma concentrations over different PK sampling times. |
| $t_{max}$ | The time at which $C_{max}$ was apparent. |
| $AUC_{0-t}$ | The area under the concentration versus time curve from time zero to the last quantifiable concentration ($C_{last}$), calculated by the linear up-log down trapezoidal method. |
| $AUC_{0-12h}$ | The area under the concentration versus time curve from time zero to 12 h post-dose, calculated by the linear up-log down trapezoidal method. |
| $AUC_{0-24h}$ | The area under the concentration versus time curve from time zero to 24 h post-dose, calculated by the linear up-log down trapezoidal method. |
| $AUC_{0-36h}$ | The area under the concentration versus time curve from time zero to 36 h post-dose, calculated by the linear up-log down trapezoidal method. |

Systemic exposure to miR29a ($AUC_{0-t}$ and $C_{max}$) in males will be compared to that in females using dose-adjusted plots.

Actual sampling times will be used for the PK analysis. Where the actual sampling time is not recorded, the nominal sampling time will be used.

All calculations will be made using raw (unrounded) data.

#### 6.3 Pharmacokinetic analysis

Plasma concentration data will be summarised by sampling time and dose level, as appropriate; PK parameters will be summarised by dose level.

All individual plasma and PK parameter estimates will be listed and summarised. Mean and individual plasma concentration versus time profiles will be illustrated using both linear-linear and logarithmic-linear scales.

Summary statistics will include number of observations (n), arithmetic mean and SD. Summaries for the PK parameters will also display the median, minimum and maximum. In addition, with the exception of $t_{max}$, the geometric mean and geometric coefficient of variation (CV)

where $CV = \sqrt{\exp(SD_{\ln}^{2}) - 1} * 100$($SD_{\ln}$ is the standard deviation of the natural logarithmically transformed data) will be reported for all PK parameters.

Between-subject variability will be based on geometric mean CVs.

Plasma concentrations and PK parameters will be reported to 3 significant figures.

##### 6.4 Assessment of dose proportionality

An equivalence approach [5, 6] will be used to assess dose proportionality. The exponents and 90 % confidence intervals (CIs), $b_{lower}$ ($b_l$) and $b_{upper}$ ($b_u$), will be presented. The estimate of the fold increase in exposure for a doubling in dose (with 90 % CI) will also be presented. The increase in exposure expected for a doubling in dose will be calculated as $2^b$ (90 % CI: $2^{bl}$, $2^{bu}$); there would be evidence of dose-proportionality if the 90 % CI lies within the limits of 1.6 to 2.5. The assessment of dose proportionality will be carried out using the Linear Mixed Effects Wizard in WinNonlin Phoenix Version 8.3 (or higher version). The assumption of a linear relationship between the $\log_e$ transformed PK parameter and $\log_e$ dose will be tested by including a quadratic term $[\log_e (\text{dose})]^2$ in the model to test for departures from linearity. The p-value from this test will be presented as a test for lack of fit (LOF) of the power model. If the LOF p-value is significant at the 5% level ($p<0.05$), an alternative method based on analysis of variance (ANOVA) will also be presented.

#### 7 DEVIATIONS FROM PROTOCOL

Deviations from the clinical protocol that are relevant to the PK analysis will be documented in the PK Report Appendix.

#### 8 INTERIM REPORT REQUIREMENTS

There are planned to be three Interim Reports using nominal blood sampling times. The Interim Report will include plasma concentrations and PK parameters of miR29a for each subject with summary statistics. Figures illustrating the individual and mean plasma concentration-time profiles will be incorporated. After the first cohort, an assessment of dose proportionality will also be included. An independent quality control review will be carried out on the analysis and reporting of each cohort.

#### 9 FINAL REPORT APPENDIX REQUIREMENTS

KinetAssist will issue a single unaudited draft report appendix for the Sponsor’s review and comments before the report is finalised.

A final report appendix will be supplied to the Sponsor and for incorporation into the clinical study report and will be archived with the Sponsor. The authorised final report appendix will be provided in Word and PDF format (paper originals of the handwritten signature pages will also be provided).

#### 10 RECORDS TO BE MAINTAINED

Records to be maintained as raw data for this work will include, where applicable, but will not be limited to the following:

Final signed protocol and any amendments
Original signed PK Plan
Original signed PAP amendments
Deviations from the PAP and or SOPs (as filenote)
Raw (received) data printouts
Computer system printouts
Tabulated and plotted data

#### 11 STORAGE OF DATA

A copy of the final report appendix and the records specified in Section 10 will be delivered to the Sponsor for the purpose of archiving. Archiving and data storage is described in KinetAssist's SOP-10 [7].

#### 12 REFERENCES

[1] Quality Control of Data Results and Reports. SOP-08.

[2] The standardisation of pharmacokinetic symbols. SOP-03.

[3] Pharmacokinetic parameters for basic study designs. SOP-04.

[4] WinNonlin Phoenix Version 8.3. Certara USA, Inc., 103 Carnegie Center, Suite 300, Princeton, NJ 08540 USA (2020).

[5] Hummel J, Mckendrick S, Brindley C, French R. Exploratory assessment of dose proportionality: Review of current approaches and proposal for a practical criterion. Pharmaceutical Statistics 2008; 8:38-49.

[6] McKendrick S, Hummel J. A Power Study for the Use of Equivalence Criteria to Assess Dose Proportionality. Poster presentation at PKUK Conference, Edinburgh, UK, November 2007.

[7] Archiving and data storage. SOP-10.

### 13 COMPUTER SYSTEMS

The following computer systems will be used throughout this work:

Microsoft Excel
Microsoft Word
WinNonlin Phoenix Version 8.3

### 14 PHARMACOKINETIC ANALYSIS PLAN DISTRIBUTION

The PAP will be distributed to the Sponsor and MAC Clinical Research Manchester (Early Phase Unit).

### 15 LIST OF TABLES AND FIGURES

The listings detailed below may be subject to alteration

#### LIST OF TABLES

Table 1 Individual and mean plasma values of miR29a (relative expression of miR29a compared to miR159a control) following a single injection of TenoMiR at 200 µg/mL to subjects

Table 2 Individual and mean plasma values of miR29a (relative expression of miR29a compared to miR159a baseline) following a single injection of TenoMiR at 200 µg/mL to subjects

Table 3 Individual and mean plasma values of miR29a (relative expression of miR29a compared to miR159a control) following a single injection of TenoMiR at 500 µg/mL to subjects

Table 4 Individual and mean plasma values of miR29a (relative expression of miR29a compared to miR159a baseline) following a single injection of TenoMiR at 500 µg/mL to subjects

Table 5 Individual and mean plasma values of miR29a (relative expression of miR29a compared to miR159a control) following a single injection of TenoMiR at 2000 µg/mL to subjects

Table 6 Individual and mean plasma values of miR29a (relative expression of miR29a compared to miR159a baseline) following a single injection of TenoMiR at 2000 µg/mL to subjects

Table 7 Individual and mean pharmacokinetic parameters of miR29a following a single injection of TenoMiR at 200 µg/mL to subjects (relative expression of miR29a compared to miR159a control)

Table 8 Individual and mean pharmacokinetic parameters of miR29a following a single injection of TenoMiR at 200 µg/mL to subjects (relative expression of miR29a compared to miR159a baseline)

Table 9 Individual and mean pharmacokinetic parameters of miR29a following a single injection of of TenoMiR at 500 µg/mL to subjects (relative expression of miR29a compared to miR159a control)

Table 10 Individual and mean pharmacokinetic parameters of miR29a following a single injection of of TenoMiR at 500 µg/mL to subjects (relative expression of miR29a compared to miR159a baseline)

Table 11 Individual and mean pharmacokinetic parameters of miR29a following a single injection of TenoMiR at 2000 µg/mL to subjects (relative expression of miR29a compared to miR159a control)

Table 12 Individual and mean pharmacokinetic parameters of miR29a following a single injection of TenoMiR at 2000 µg/mL to subjects (relative expression of miR29a compared to miR159a baseline)

Table 13 Relationship between $C_{max}$ and $AUC_{0-t}$ values of miR29a and dose of TenoMiR following a single injection of TenoMiR to subjects (relative expression of miR29a compared to miR159a control)

Table 14 Relationship between $C_{max}$ and $AUC_{0-t}$ values of miR29a and dose of TenoMiR following a single injection of TenoMiR to subjects (relative expression of miR29a compared to miR159a baseline)

Table 15 Actual blood sampling times

Table 16 Subject's age, sex and bodyweight

#### LIST OF FIGURES

Figure 1 Mean plasma values of miR29a (relative expression of miR29a compared to miR159a control) following a single injection of TenoMiR at 200, 500 and 2000 µg/mL to subjects

Figure 2 Mean plasma values of miR29a (relative expression of miR29a compared to baseline) following a single injection of TenoMiR at 200, 500 and 2000 µg/mL to subjects

Figure 3 Individual plasma values of miR29a (relative expression of miR29a compared to miR159a control) following a single injectionof TenoMiR at 200 µg/mL to subjects

Figure 4 Individual plasma values of miR29a (relative expression of miR29a compared to baseline) following a single injection of TenoMiR at 200 µg/mL to subjects

Figure 5 Individual plasma values of miR29a (relative expression of miR29a compared to miR159a control) following a single injection of TenoMiR at 500 µg/mL to subjects

Figure 6 Individual plasma values of miR29a (relative expression of miR29a compared to miR159a control) following a single injection of TenoMiR at 500 µg/mL to subjects

Figure 7 Individual plasma values of miR29a (relative expression of miR29a compared to miR159a control) following a single injection of TenoMiR at 2000 µg/mL to subjects

Figure 8 Individual plasma values of miR29a (relative expression of miR29a compared to miR159a control) following a single injection of TenoMiR at 2000 µg/mL to subjects

Figure 9 Relationship between $C_{max}$ and $AUC_{0-t}$ values of miR29a and dose of TenoMiR following a single injection of TenoMiR to subjects (relative expression of miR29a compared to miR159a control)

Figure 10 Relationship between $C_{max}$ and $AUC_{0-t}$ values of miR29a and dose of TenoMiR following a single injection of TenoMiR to subjects (relative expression of miR29a compared to baseline)

Figure 11 Dose-adjusted $C_{max}$ and $AUC_{0-t}$ values of TenoMiR following single inhaled doses of 5 mg TenoMiR to male and female subjects (relative expression of miR29a compared to miR159a control)

Figure 12 Dose-adjusted $C_{max}$ and $AUC_{0-t}$ values of TenoMiR following single inhaled doses of 5 mg TenoMiR to male and female subjects (relative expression of miR29a compared to baseline)
