## Supplementary material for "First in Human study of a microRNA29a mimic (TenoMiR) in patients with lateral elbow tendinopathy - a randomised, Placebo Controlled Phase 1 trial": Statistical analysis plan

A Phase 1, Single-Centre, Randomised, Double-Blind, Placebo-Controlled Study Evaluating the Safety, Tolerability and Pharmacokinetics of Single Ascending Doses of TenoMiR Injections in Subjects with Lateral Epicondylitis

Investigational Medicinal Product: TenoMiR

Study Code: CWT-TE1

MAC STUDY CODE: MAC075

Version: 1.0
date: 22FEB21

**Issued by:**

**Paul Steven, MAC Clinical Research, Statistician**

Signature:	Date:

**Approved by (Sponsor):**

**Dr Derek Gilchrist, Causeway Therapeutics**

Signature:	Date:

LIST OF ABBREVIATIONS AND DEFINITIONS OF TERMS

The following abbreviations and special terms are used in this document.

| **Abbreviation** | **Explanation** |
| --- | --- |
| AE | Adverse Event |
| ASES-E | American Shoulder and Elbow Surgeons Elbow |
| AUC | Area under the curve |
| BP | Blood Pressure |
| eCRF | Electronic Case Report Form |
| CI | Confidence Intervals |
| C_max_ | Maximum Concentration |
| DBP | Diastolic Blood Pressure |
| ECG | Electrocardiogram |
| GLMM | Generalised Linear Mixed Model |
| HR | Heart Rate |
| MedDRA | Medical Dictionary for Regulatory Activities |
| PK | Pharmacokinetics |
| PRTEE | Patient-Rated Tennis Elbow Evaluation |
| PRO | Patient reported outcome |
| PT | Preferred Term |
| RR | Respiratory Rate |
| SAE | Serious Adverse Event |
| SAS | Statistical Analysis system |
| SBP | Systolic Blood Pressure |
| SRC | Safety Review Committee |
| SOC | System Organ Class |
| SD | Standard Deviation |
| TEAEs | Treatment-Emergent Adverse Events |
| T_max_ | Time taken to reach C_max_ |
| QuickDASH | The Disabilities of the Arm, Shoulder and Hand Score |

1 Study Details
  1.1 INTRODUCTION

The purpose of this Statistical Analysis Plan (SAP) is to define the efficacy and safety analysis variables and analysis methodology to address the study objectives.

The PK analysis is outside the scope of this analysis plan. That analysis will be described in a separate document.

The protocol dated 23 July 2020 (version 3.0) was used in the development of this statistical analysis plan.

1.2 STUDY OBJECTIVES

The objectives of the study are:

Primary Objective

The primary objective of the study is to determine the safety and tolerability of single ascending doses of TenoMiR in subjects with lateral epicondylitis.

Secondary Objective

The secondary objectives of the study are:

- To determine the single dose pharmacokinetics (PK) of TenoMiR administration in subjects with lateral epicondylitis.
- To assess the efficacy of TenoMiR administration in subjects with lateral epicondylitis.
  1.3 STUDY ENDPOINTS

**Primary Endpoint**

The primary endpoint of the study is the comparison of safety data between TenoMiR versus placebo as measured by incidence of AEs, clinical laboratory abnormalities, changes in vital signs (blood pressure, temperature, respiratory rate, and pulse rate), 12-lead ECG parameters, physical examinations and skin score assessment at 14 days post injection.

**Secondary Endpoints**

The secondary endpoints of the study are:

- The plasma PK of TenoMiR as shown by maximum drug plasma concentration (C_max_), time to reach C_max_ (t_max_) and area under the plasma vs. concentration time curve (AUC).
- Efficacy of a single dose of TenoMiR on elbow pain as measured using Visual Analogue Scale (VAS) from pre-dose on Day 1 to Day 14, Day 28 and Day 90.
- Efficacy of a single dose of TenoMiR on disability and symptoms measured using the Disabilities of the Arm, Shoulder, and Hand (Quick DASH) Score from pre-dose on Day 1 to Day 14, Day 28 and Day 90.
- Efficacy of a single dose of TenoMiR on pain and disability as measured by the American Shoulder and Elbow Surgeons Elbow (ASES-E) Score from pre-dose on Day 1 to Day 14, Day 28 and Day 90.
- Efficacy of a single dose of TenoMiR on pain and disability as measured by the Patient Rated Tennis Elbow Evaluation (PRTEE) from pre-dose on Day 1 to Day 14, Day 28 and Day 90.
- Efficacy of a single dose of TenoMiR on lateral elbow tendons as measured by change from baseline ultrasound assessment from pre-dose on Day 1 to Day 28 and Day 90
  1.4 STUDY DESIGN

This is a Phase 1, randomised, double-blind, placebo-controlled study evaluating the safety, tolerability and PK of single ascending doses of TenoMiR injections in subjects with lateral epicondylitis (see Figure 1 for study design).

It is planned to enrol between 24 and 32 subjects (pending dose evaluation) with lateral epicondylitis.

Subjects will be divided into cohorts (8 subjects per cohort) and will be randomised (3:1) to receive a single 1 mL dose of TenoMiR or saline placebo (0.9%) administered via injection. The starting dose of TenoMiR will be 200 µg/mL. The doses will be escalated to 500 µg/mL and then 1500 µg/mL. All doses will be given as a 1mL injection under ultrasound guidance into the affected area. One additional cohort (8 subjects) may be utilised to explore an additional dose, if necessary. The decision to explore an additional dose will be made by an interim analysis group comprising the Sponsor and Safety Review Committee (SRC). In each cohort, no more than 2 subjects on the first dosing day (1 active; 1 placebo) will be dosed, such that no more than 1 subject will receive an active TenoMiR dose for the first time at each dose level. Depending on the safety and tolerability of the previously dosed sentinel subjects, dosing will continue in the remaining subjects (6 subjects; 5 active, 1 placebo) at the same dose level in each cohort. After 72 hours, depending on the safety and tolerability of the previously dosed sentinel subjects, dosing will continue in the remaining subjects (6 subjects; 5 active, 1 placebo) at the same dose level in each cohort.

Doses will be administered in an escalating manner, following satisfactory review of all safety, tolerability, plasma PK and efficacy data (where available) from lower doses. Except for the starting dose, the doses outlined in the Protocol are preliminary, with actual subsequent doses determined based on the ongoing evaluation of the safety, tolerability, plasma PK and efficacy data (where available) by the SRC.

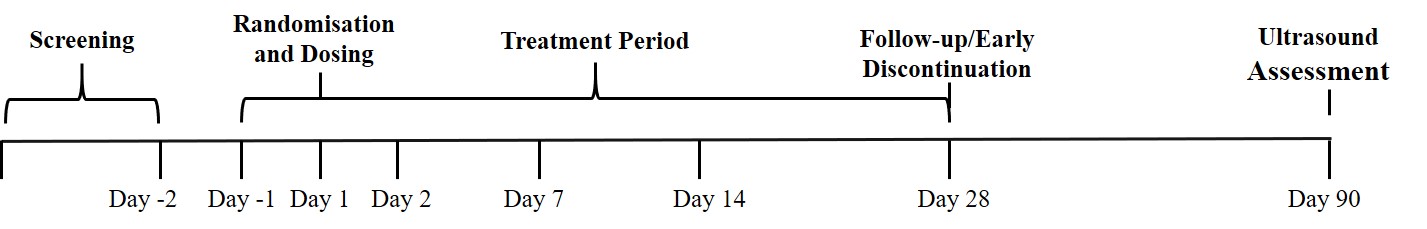

Figure 1: Study Design

2 ANALYSIS SETS
  2.1 Efficacy Analysis Set (Per protocol)

Every randomised patient who is treated with TenoMiR or placebo and contributes at least 1 post-dose result for any elbow rating scale will be included in the Efficacy Analysis Set (EAS). This population will be used for all efficacy analyses and efficacy analysis will be performed by actual treatment received.

2.2 Safety Analysis Set

Every randomised patient who is treated with TenoMiR will be included in the Safety Analysis Set. Safety tables will be presented by actual treatment received.

2.3 Pharmacokinetic Analysis Set

The PK Population is defined as all subjects who received a dose of TenoMiR on Day 1 and have evaluable plasma concentrations of miR29a.

2.4 Protocol Deviations

Protocol deviations will be categorized as minor/major and deviation category in a blinded manner prior to the database lock for each study part. The sponsor will review protocol deviations throughout the course of the study and provide the classifications for each deviation. These will be summarized by Major/Minor and deviation category. All deviations will be listed.

3 Primary and Secondary Variables

Patient reported outcome (PRO) questionnaires will be assessed using the Pain VAS, ASES-E, PRTEE, and QuickDASH. All items/questionnaires will be scored according to published scoring guidelines or the developer’s guidelines, if published guidelines are not available. All PRO analyses will be based on the FAS, unless otherwise stated.

3.1 Pain Visual Analogue Scale

The Pain Visual Analogue Scale is reported as a numeric integer between 0 and 100. For all efficacy modelling and descriptive summaries, the Pain VAS will be reported as per recorded, with no further derivations required.

3.2 ASES-E Score

The ASES-E Score contains 3 Patient Self-Evaluation scales and 4 Physician assessment scales.

The sub sub-scales are

- Patient Self Evaluation: Pain
  - 5 items, ordered scale 1-10
- Patient Self Evaluation: Function
  - 12 items per arm, ordered scale 0-3
- Patient Self Evaluation: Satisfaction
  - 1 items, ordered scale 1-10
- Physician Assessment: Motion
  - 5 items, continuous response: degrees of motion
- Physician Assessment: Stability
  - 3 items per arm, ordered scale 0-3
- Physician Assessment: Strength
  - 5 items per arm, ordered scale 0-5
- Physician Assessment: Signs
  - 8 items per arm, ordered scale 0-3
  - 14 items per arm, Y/N response

For each of the following sub scales, the sum of the scores for each item will be added to create an overall score.

- Patient Self Evaluation: Pain
  - Add up 5 items. Best score= 0; Worst score = 50
- Patient Self Evaluation: Function
  - Add up 12 items, using reversed responses (e.g. score = 3 – original score). Best score= 0; Worst score = 36
- Patient Self Evaluation: Satisfaction
  - Single item, best score = 10, worst score = 1
- Physician Assessment: Motion
  - Scores are assessed separately and not combined.
- Physician Assessment: Stability
  - Add up 3 items. Best score= 0; Worst score = 9
- Physician Assessment: Strength
  - Add up 5 items, using reversed responses (e.g. score = [5 – original score]). Best score= 0; Worst score = 25
- Physician Assessment: Signs
  - Yes/no questions are summarised descriptively
3.3 Patient Rated Tennis Elbow Evaluation (PRTEE)

The PRTEE contains 2 sub-scales; Pain and Function. Higher score indicates more pain and functional disability (e.g., 0 = no disability). The details are as follows:

- Pain subscale, 0 = no pain, 10 = worst imaginable
  - 5 items
- Function subscale, 0 = no difficulty, 10 = unable to do
  - Specific activities
    - 6 items
  - Usual activities
    - 4 items

Totals are calculated for each individual subscale. An overall total score is computed by weighting equally each of the two subscales, to give an overall score out of 100.

The totals are derived as follows:

- Pain subscale Total

Add up 5 items. Best score= 0; Worst score = 50

- Function: Specific Activities Total

Add up 6 items Best Score= 0; Worst Score = 60

- Function: Usual Activities Total

Add up 4 items items Best Score= 0; Worst Score = 40

- Function subscale Total

(Total Specific Activities + Total Usual Activities)/2

Best score= 0; Worst score = 50

- Total PRTEE Score =

Pain Subscale + Function Subscale

Best Score= 0 Worst Score = 100

(pain and disability contribute equally to score)

The actual change from baseline in PRTEE score will be derived for each visit where there is available data. For example; at visit X, the calculation will be (PRTEE score at visit X – baseline PRTEE score). Actual change from baseline for the individual PRTEE subscale scores and the total will be calculated in a similar way. The baseline score is defined as last non-missing score prior to dosing with study treatment. Any questionnaires completed on Day 1 of dosing will be considered pre-dose.

3.4 Quick Disability of the Arm, Shoulder, and Hand (DASH) Score

The QuickDASH contains a single scale which gives a Disability / Symptom score. The details are as follows:

- Disability / Symptoms (1 = no difficulty, 5 = unable to do)
  - 11 items
- Work Module (optional) (1 = no difficulty, 5 = unable to do)
  - 4 items
- Sports / Performing Arts Module (optional) (1 = no difficulty, 5 = unable to do)

The QuickDASH Disability / Symptoms total score is calculated using the following formula

Total score = [(Sum of n responses / n) – 1] x 25

Where n is equal to the number of completed responses.

For the optional modules, a total score for each module is calculated using the following formula:

Total score = [(Sum of responses / 4) – 1] x 25

The QuickDASH Disability / Symptoms total score is not calculated if there is greater than 1 missing item. An optional module score is not calculated if there are any missing items.

3.5 Missing data

For the ASES-E and PRTEE, if there are missing items, subscale scores can be prorated. This can be done on the scoring guide or by using the formula below:

Prorated subscale score = ([Sum of item scores] × [N of items in subscale]) / [N of items answered]

When there are missing data, prorating by subscale in this way is acceptable as long as more than 50% of the items were answered (a minimum of 4 of 7 items, 4 of 6 items, etc.). The total score is then calculated as the sum of the unweighted subscale scores.

4 Analysis Methods
  4.1 General Principles

All summary tables, figures and data listings will be produced using SAS software 9.4 or above.

Summaries will be performed by treatment group and overall.

Descriptive statistics will be used for all variables, as appropriate. Continuous variables will be summarised by the number of observations, mean, standard deviation, median, minimum, and maximum. For log-transformed data, the geometric mean, coefficient of variation (CV), median, minimum and maximum will be presented.

Categorical variables will be summarised by frequency counts and percentages for each category.

For adverse event (AE) summaries, the n and % of patients with event, as well as the number of events, will be shown.

In general, for efficacy and PRO endpoints the last observed measurement prior to randomization will be considered the baseline measurement. However, if an evaluable assessment is only available after randomization but before the first dose of randomized treatment then this assessment will be used as baseline. For safety endpoints the last observation before the first dose of study treatment will be considered the baseline measurement unless otherwise specified. For assessments on the day of first dose where time is not captured, a nominal pre-dose indicator, if available, will serve as sufficient evidence that the assessment occurred prior to first dose.

Assessments on the day of the first dose where neither time nor a nominal pre-dose indicator are captured will be considered prior to the first dose if such procedures are required by the protocol to be conducted before the first dose.

In all summaries change from baseline variables will be calculated as the post-treatment value minus the value at baseline. The percentage change from baseline will be calculated as (post-baseline value - baseline value) / baseline value x 100.

All listings will be ordered by treatment group (TenorMiR or placebo) and patient number, and will include all available data including unscheduled data.

Unscheduled values will be mapped to the closest prior nominal visit/timepoint. If multiple values exist for a nominal visit/timepoint then the worst value of available results will be used in the summary.

Listings will show the original visit and timepoint as collected on the CRF as well as the mapped visit and timepoint for unscheduled values. An indicator will be included in the listings to indicate which value is utilized in the analysis.

4.2 Patient Accountability

Summaries of analysis populations and patient disposition will be summarized and listed using all patients who have signed informed consent. The data summaries will contain the following information:

- Number of patients randomized
- Number and percent of patients who received TenorMiR/placebo
- Number and percent of patients who completed the study
- Number and percent of patients who discontinued early from the study and reason for early discontinuation
- Number and percent of patients in each of the analysis populations
  4.3 DEMOGRAPHICS AND BASELINE CHARACTERISTICS

The full analysis (efficacy) set population will be used for all demographic and baseline characteristics summaries. All demographics and baseline characteristics data will be included in listings. The actual treatment (TenorMiR / placebo) received will be utilized in these analyses.

4.3.1 Demographics

Demographics including age, ethnicity, height, weight, gender, body mass index will be summarised appropriately. In general, for the continuous demographic variables (i.e., age, height, weight and body mass index) results for each treatment group will be summarised using mean, SD, median, and minimum and maximum values. For categorical (nominal) variables (i.e., race, ethnicity, and gender), the number and percentage of patients will be used.

4.3.2 Medical History

Medical history will be summarised by treatment group using number of observations and percentages of patients reporting each category. Medical history will be coded into the most recent version of the Medical Dictionary for Regulatory Activities (MedDRA) available when coding activities commence.

4.3.3 Eligibility Laboratory Assessments

Pregnancy test, follicle stimulating hormone test, urine cotinine, urine drugs of abuse, alcohol breath screen, SARS-CoV-2 test and serology will be listed by patient, treatment, and timepoint, as appropriate.

4.4 PRIOR/CONCOMITANT MEDICATIONS AND RESCUE MEDICATIONS

All prior or concomitant and rescue medication data will be included in listings.

Medications will be reported using the most recent version of the World Health Organization drug (WHODRUG) codes available when coding activities commence. Medications will be classified according to the World Health Organization Drug Dictionary (WHODD Version Mar2019 or later) ATC code levels 2 to 4.

Prior medications are defined as medications taken prior to the dosing of TenorMiR/placebo.

Concomitant medications are defined as medications taken on or after the date/time of TenorMiR/placebo regardless of the start date/time. Medications can be classified as both prior and concomitant. Prior and concomitant medications will also be listed by ATC code level 4. Actual treatment received will be utilized in summaries.

Rescue medication is defined as any medication utilized for pain relief after dosing of TenorMiR/placebo. These medications will be summarized by ATC code level 2 and preferred term (PT). In addition, number of rescue medication doses will be summarized. Actual treatment will be utilized in summaries.

4.5 Efficacy Assessments

The analysis of efficacy will be descriptively summarized overall and by treatment and timepoint, as appropriate. NRS scores will be graphed by treatment and timepoint.

Data will be presented and compared by actual treatment but may be analyzed by randomized treatment in a supplementary manner if deemed necessary. Patients who received IP other than the IP they were randomized to will be noted in the listings.

Table 1 details which efficacy endpoints are to be analysed, together with details of the analysis methods.

**Table 1: Planned statistical analysis of efficacy parameters**

| **Endpoint analysed** | **Notes** |
| --- | --- |
| Elbow pain as measured using Visual Analogue Scale (VAS) from pre-dose on Day 1 to Day 14, Day 28 and Day 90.  Disability and symptoms measured using the Disabilities of the Arm, Shoulder, and Hand (Quick DASH) Score from pre-dose on Day 1 to Day 14, Day 28 and Day 90.  Disability and symptoms measured using the American Shoulder and Elbow Surgeons Elbow (ASES-E) Score from pre-dose on Day 1 to Day 14, Day 28 and Day 90.  Pain and disability as measured by the Patient Rated Tennis Elbow Evaluation (PRTEE) from pre-dose on Day 1 to Day 14, Day 28 and Day 290.  Lateral elbow tendons as measured by change from baseline ultrasound assessment from pre-dose on Day 1 to Day 28 and Day 90. | **Secondary**: Differences in scores for each timepoint  **Analysis**: GLMM with efficacy parameter as response variable, treatment, timepoint and interaction between treatment and timepoint as fixed effects, baseline measure as a covariate and patient and elbow (left and right) as random effects. Treatment difference between active and placebo at each timepoint will be reported, along with corresponding 95% confidence intervals  **For continuous data**: Treatment difference reported as differences in least squares mean  **For categorical data**: Treatment difference reported as odds ratio |

4.5.1 Efficacy analysis

Descriptive statistics and modelling will be performed for the following scales and sub-scales.

Pain VAS:

Item Score

QuickDASH:

QuickDASH Total Score

QuickDASH optional scales

ASES-E:

Patient Self Evaluation: Pain Total Score

Patient Self Evaluation: Function Total Score

Patient Self Evaluation: Satisfaction Total Score

Physician Assessment: Motion

Physician Assessment: Strength

PRTEE:

Pain subscale Total

Function: Specific Activities Total

Function: Usual Activities Total

Function subscale Total

PRTEE Score: Total

Additionally, summaries will be produced on an individual item basis for each scale.

Descriptive statistics

Descriptive statistics and graphs will be reported for the following scales and subscales by visits as well as change in these scores from baseline. All items, scale, and sub-scale totals will be listed.

Modelling

Change from baseline in score will be regarded as the main efficacy analysis for each of the VAS Pain scale, QuickDASH assessment, ASES-E assessment and PRTEE assessment. It will be analysed using a Generalized linear mixed model (GLMM) analysis of the change from baseline (defined as on or prior to first dose) in score for each visit.

A GLMM will be fit to the data with efficacy parameter as response variable, treatment, timepoint and interaction between treatment and timepoint as fixed effects, baseline measure as a covariate and patient and elbow (left and right) as random effects.

For each treatment and visit, the adjusted (least squares) mean estimates, corresponding 95% CIs, estimates of the treatment difference and corresponding 95% CIs will be presented.

4.5.2 Ultrasound Assessment

Analysis of the ultrasound assessments is outside of the scope of this SAP and will be analysed separately.

4.6 Safety Parameters

The objective of the evaluation of the safety variables is to investigate the data for any effects on clinical tolerability and laboratory safety variables.

Safety assessments include AE monitoring, standard laboratory safety evaluations (haematology, blood chemistry and urinalysis), supine vital signs (RR, BP, HR and oral temperature), orthostatic vital signs, physical examinations, 12-lead ECG and skin score assessments.

The analysis of safety will be descriptively summarized by timepoint, as appropriate. Data will be presented by actual treatment received for the purposes of summarizing the safety results.

No formal hypothesis testing will be carried out.

4.6.1 Adverse Events

Adverse events will be listed in the following categories:

- Treatment-emergent adverse events (TEAEs) is any adverse event with a start date and time on or after dosing with TenoMiR/placebo. All TEAEs will be listed and summarized as indicated below

The overall incidence of TEAEs (number and percentage of patients) as well as the number of events will be summarized by treatment and overall, categories of degree of severity, SAEs, causally related TEAEs and SAEs, TEAEs leading to discontinuation of treatment and AEs or SAEs leading to withdrawal.

TEAEs will be summarized at both the patient [number (%) of patients] and event [number of events] level for the following:

- SOC and PT
- SOC and PT and maximum reported severity
- SOC and PT and causal relationship to study drug

These analyses will be repeated for SAEs and treatment related AE if deemed necessary, otherwise only the SAEs by SOC and PT will be performed.

For the incidence at the patient level by SOC and PT; if a patient experiences more than one event within the same SOC and PT, only one occurrence will be included in the incidence.

For the incidence at the patient level by SOC, PT and severity; if a patient experiences more than one event within the same SOC and PT, only the most severe occurrence will be included in the incidence.

4.6.2 Clinical Laboratory Evaluations

Clinical laboratory evaluation results (chemistry and haematology) will be listed for individual patients and compared to laboratory reference ranges and those values outside of the applicable range will be flagged as high (H) or low (L) and categorised by clinical significance (clinically significant/non-clinically significant). This classification of high, low and normal will be summarized by panel (i.e., chemistry and haematology), laboratory test, visit and timepoint.

For all laboratory variables, baseline value will be calculated as last laboratory value prior to dosing. Change from baseline values at each assessment will be calculated as the assessment value minus the baseline value. The quantitative laboratory data, along with changes from baseline will be summarised using descriptive statistics by timepoint.

Urinalysis data will be listed only.

4.6.3 Vital Signs

Vital signs data and body temperature will be listed for individual patients.

Vital signs include heart rate (HR), systolic blood pressure (SBP), diastolic blood pressure (DBP), respiration rate and body temperature.

Vital signs data along with changes from baseline and percent change from baseline will be summarised using descriptive statistics by position and timepoint. For each vital sign variable, baseline value will be calculated from the mean of the available triplicate (if collected) pre-dose values.

4.6.4 Electrocardiogram (ECG)

Standard 12-lead ECG parameters (PR, RR, QRS, QT, QTcF and HR) and ECG abnormal assessment will be listed for individual patients.

For each ECG variable, baseline value will be calculated from the means of the available data prior to dose, if multiple values are recorded. Change from baseline values will be calculated as the assessment values minus baseline values.

Descriptive statistics will be calculated for absolute value of each parameter, together with the corresponding changes from baseline overall and by dose and timepoint. Where multiple values are recorded at a timepoint for a patient, the mean of the available values will be used in the summary statistics.

4.6.5 Physical Examination

Physical examination data will be listed only.

4.6.6 Skin Score Assessment

Skin score assessment data will be listed only.

4.7 Safety Review Committee

For each SRC, safety and PK data will be summarised. The PK analysis required for the SRC meetings will be detailed within the PK analysis plan and is outside of the scope of this SAP. For safety, the following data will be listed only: Adverse Events, Vital Signs, ECG and Physical Examination.

5 HANDLING OF MISSING OR INCOMPLETE DATA

Unrecorded values will be treated as missing. Efforts will be made to avoid this from occurring. For complete missing visits the missing efficacy, safety will be treated as missing at random and will not be imputed in the statistical analysis except for partial dates.

For missing single items for PROMs, the imputation detailed within Section will be followed.

5.1 Prior and Concomitant Medication Dates

Partial dates for any prior and concomitant medications recorded in the eCRF will be imputed using the following convention:

- If the partial date is a start date, a ‘01’ will be used for the day and ‘Jan’ will be used for the month.
- If the partial date is a stop date, a ‘28/29/30/31’ will be used for the day (dependent on the month and year) and ‘Dec’ will be used for the month.

The recorded partial date will be displayed in listings. No imputation will be performed for completely missing start and end dates.

5.2 AE Start and End Dates

The eCRF allows for the possibility of partial dates (i.e. only month and year) to be recorded for AE start and end dates; that is the day of the month may be missing. In such a case, the following conventions will be applied for calculating the time of onset and the duration of the event:

- Missing Start Day: First of the month will be used unless this is before the start date of first study treatment; in this case the study treatment start date will be used and hence the event is considered treatment emergent.
- Missing Stop Day: Last day of the month will be used, unless this is after the stop date of study completion; in this case the study stop date will be used.

Completely missing start or end dates will remain missing, with no imputation applied. Consequently, time to onset and duration of such events will be missing.

6 TABLE AND LISTING SHELLS

TFL Shells will be provided as a separate document. The TFLs listed and the corresponding numbering may be subject to alteration. Tables and figures may be presented as in-text tables and figures in the CSR body.
