## Supplementary Figures1 & 2 for "First in Human study of a microRNA29a mimic (TenoMiR) in patients with lateral elbow tendinopathy - a randomised, Placebo Controlled Phase 1 trial"

Supplementary Figure 1

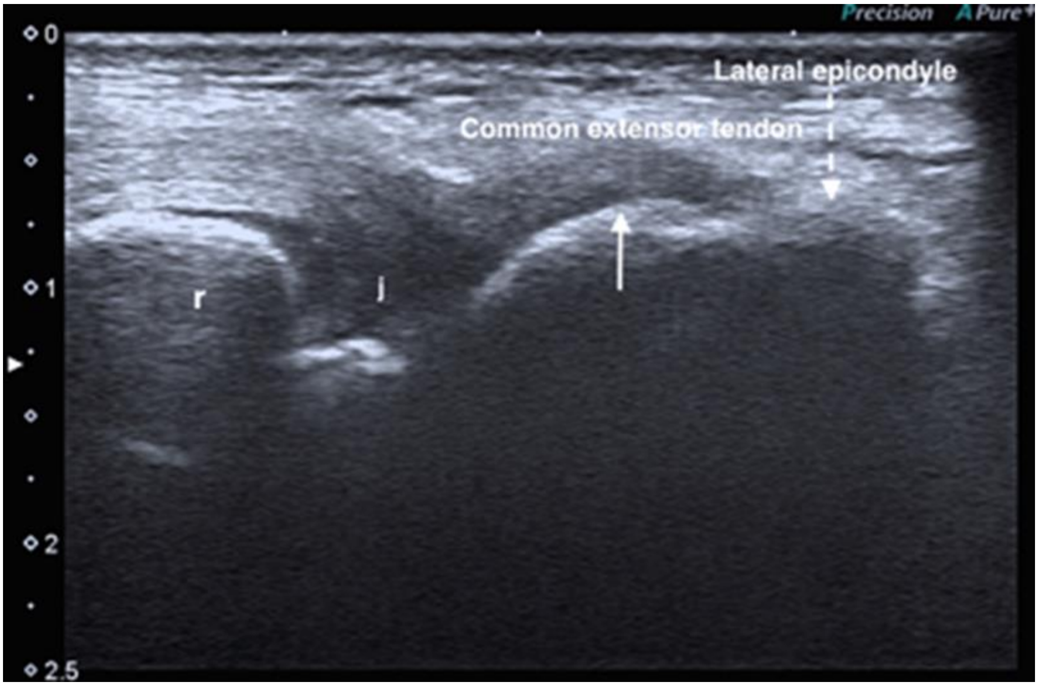


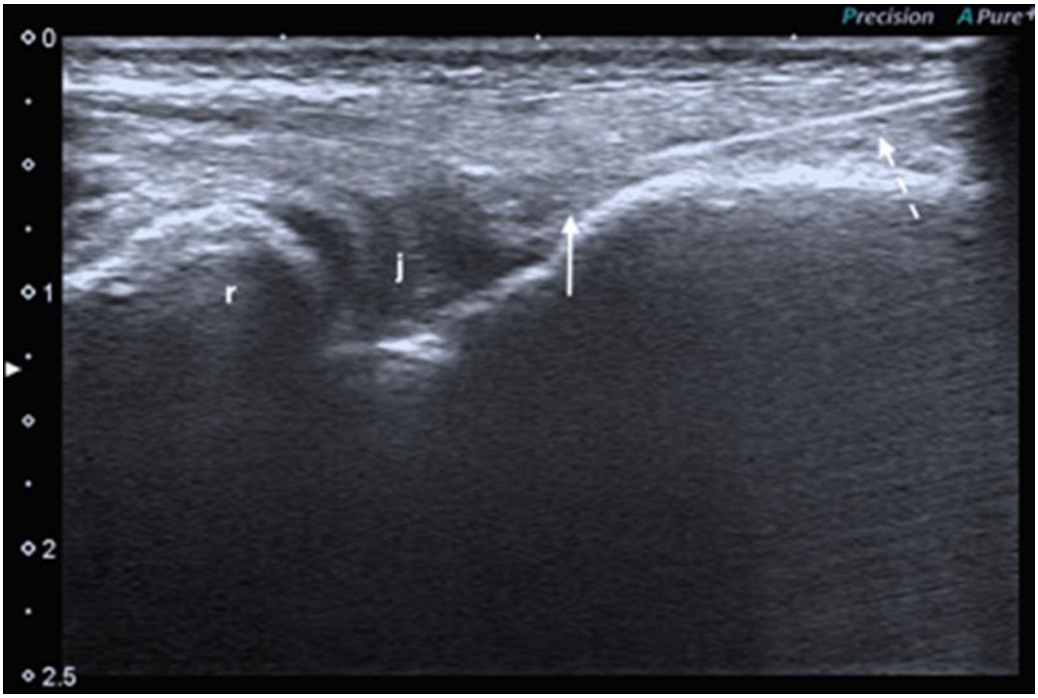

| Schedule of Assessments | Screening | | Treatment Period | | | | FU | EOS |
|---|---|---|---|---|---|---|---|---|
| | -4 Weeks | -1 Week | Week 1 | | | Week 2 | Week 4[a] | Week 13 |
| | Day -28 to Day -3 | Day -2/ Day -1 | Day 1 | Day 2 | Day 7 (±1 day) | Day 14 (±2 days) | Day 28 (±2 days) | Day 90 (±2 days) |
| **Informed Consent** | X | | | | | | | |
| **Demography** | X | | | | | | | |
| **Inclusion/Exclusion Criteria** | X | X | X | | | | | |
| **Medical History[b]** | X | | | | | | | |
| **Urine Drugs of Abuse, Urine Cotinine and Breath Alcohol Screen[c]** | X | X | X | | | | | |
| **SARS-CoV-2 Test** | X | X | | | | | | |
| **Urine Pregnancy Test** | X | | X | | | | | |
| **FSH Test[d]** | X | | | | | | | |
| **Serology** | X | | | | | | | |
| **Randomisation** | | | X | | | | | |
| **TenoMiR/Placebo Administration** | | | X | | | | | |
| **Study Residency** | | | | | | | | |
| **Non-Residential Visit** | X | X | X[e] | X | X | X | X | X |
| **Safety Assessments[f]** | | | | | | | | |
| **Physical Examination (including height and weight)[g]** | X | | X | | X | X | X | |
| **Vital Signs (pulse rate, blood pressure and respiratory rate)[h]** | X | | *Predose, 0.5, 1, 2, 4, 8 and 12 hours post-dose* | | X | X | | |
| **Oral/tympanic/thermal body temperature** | X | X | *Predose, 0.5, 1, 2, 4, 8 and 12 hours post-dose* | X | X | X | X | X |
| **12-Lead ECG[i]** | X | | *Predose, 1, 2, 4, 8 and 12 hours post-dose* | | X | X | | |
| **Clinical Laboratory Evaluations** | X | X | | X | X | | (X)[j] | X |
| **Skin Score Assessment (erythema, pain, tenderness and swelling)** | | | *Predose, 1, 2, 3, 4, 6, 8 and 12 hours post-dose* | X | X | X | | |
| **AE Monitoring** | | | | X | | | | |
| **Concomitant Medication Monitoring[k]** | | | X | | | | | |
| **Pharmacokinetic Assessments** | | | | | | | | |
| **Blood Sampling[l]** | | | *Predose, 0.5, 1, 2, 3, 4, 6, 8 and 12 hours postdose* | *24 and 36 hours post-dose* | X | | | |
| **Efficacy Assessments** | | | | | | | | |
| **Elbow Pain/Tendon Pain Questionnaire (VAS)** | | | X[m] | | | X | X | X |
| **Quick Disabilities of the Arm, Shoulder and Hand Questionnaire (DASH)** | | | X[m] | | | X | X | X |
| **American Shoulder and Elbow Surgeons Elbow (ASES-E)** | | | X[m] | | | X | X | X |
| **Patient Rated Tennis Elbow Evaluation (PRTEE)** | | | X[m] | | | X | X | X |
| **Ultrasound Assessment of Lateral Elbow Tendons** | X[n] | | | | | | X | X |

**Schedule of Assessments**

Abbreviations: AE – adverse event; CRU – Clinical Research Unit; ECG – electrocardiogram; eCRF – electronic case report form; EOS – end of study; FSH – follicle stimulating hormone; FU – follow-up; hr – hour; min – minute; PK – pharmacokinetic.

a The follow-up visit took place 28 days after dosing or at early discontinuation (if applicable).

b Medical history recorded which of the subject's elbows was affected by lateral epicondylitis and which was their dominant hand.

c Positive drugs of abuse screen allowed for opiates allowed at Screening, but not on Day -2/Day -1 or Day 1.

d If required to show menopausal status of subject.

e Subjects were allowed to remain at the CRU overnight at the discretion of the Investigator on Day 1, if required for safety reasons.

f For timepoints with multiple assessments the assessments were conducted in this order: 12-lead ECGs, vital signs then PK blood draw. The PK sample were collected at the scheduled time post-dose.

g Height and weight measured at Screening only.

h Triplicate ECG recorded at Screening. Single readings were collected at all other timepoints.

i Predose vital signs were collected within 30 minutes predose.
